# B cell tolerance checkpoint function in multiple sclerosis and transient CD52 depletion

**DOI:** 10.1101/2025.05.07.25327140

**Authors:** Anastasia Alexaki, Fotis Baltoumas, Dimitrios Tzanetakos, Chrysoula Zografou, Theodoros Dame, Chrysoula Michaletou, Galini Kyriakaki, Aglaia G. Vakrakou, Maria Anagnostouli, Georgia Karadima, Marianna Tzanoudaki, Marinos C. Dalakas, Leonidas Stefanis, Konstantinos Kilidireas, Kevin C. O’Connor, Georgios Pavlopoulos, Panos Stathopoulos

## Abstract

Transient CD52 immune cell depletion with the monoclonal antibody alemtuzumab is highly effective in treating relapsing-remitting multiple sclerosis but often leads to secondary autoimmunity. Whether these effects are linked to an alteration of B cell tolerance mechanisms is currently not known. To evaluate peripheral B cell tolerance checkpoint integrity in patients and controls, we constructed 138 recombinant mAbs from single mature naïve B cells and tested their poly- and autoreactivity. We examined three healthy donors (HDs), three immunotherapy-naïve MS patients, and six patients treated with alemtuzumab at comparable time points post-treatment (mean ± SD 3.8 ± 0.39 years). Moreover, we investigated B cell receptor (BCR) repertoire parameters associated with tolerance mechanisms in the same subject groups. Polyreactive and autoreactive fraction means did not differ significantly among the three subgroups. Presence of a high or low autoreactive fraction of naïve B cells in patients treated with alemtuzumab did not correlate with secondary autoimmunity at the time of sampling and with future MS activity, and therefore most likely reflects stochastic variation in the context of immune reconstitution. In BCR repertoire analysis, alemtuzumab-treated patients showed lower mean naïve complementarity-determining region 3 (CDR3) net charge compared to HDs (P=0.0036), an interesting yet isolated finding warranting further investigation. Overall, transient CD52 depletion did not affect major changes in peripheral B cell tolerance checkpoint function as assessed with naïve B cell cloning and BCR NGS, while observations in the described setting may also apply to other immune reconstitution strategies.

**Highlights:**

- Naive B cell polyreactivity of aCD52-treated patients did not differ from controls.
- Naive B cell autoreactivity of a CD52-treated patients did not differ from controls.
- Naive B cell autoreactvity of aCD52-treated patients was variable.
- Variability of autoreactivity did not correlate with clinical outcomes.
- aCD52-treated patients CDR3 properties showed isolated differences from controls.

## 1. Introduction

Multiple sclerosis (MS), the most common autoimmune disease of the central nervous system (Leray et al., 2016), is pathologically characterized by demyelination, acute and chronic inflammation, and axonal and neuronal degeneration (Pitt et al., 2022). MS is associated with genetic and non-genetic risk factors (Alfredsson and Olsson, 2019; International Multiple Sclerosis Genetics Consortium, 2019), and, from a clinical standpoint, has been historically classified into subtypes, the more common being relapsing-remitting MS (RRMS) (Ntranos and Lublin, 2016).

The high clinical efficacy of anti-CD20 therapies has highlighted the crucial role of B cells in disease pathogenesis (Hauser et al., 2017). Physiologically, as B cells mature in the bone marrow, V(D)J gene recombination generates the necessary B cell receptor (BCR) diversity to shield against infection but at the same time also autoreactive BCRs. These are removed by two tolerance checkpoints: the central, between the pre-B to immature B cell transition, and the peripheral, between the new emigrant to mature naive B cell transition. Defects in checkpoint function have been found in almost all autoimmune diseases examined (Meffre and O’Connor, 2019). In RRMS, a defective central (in 2 out of 7 patients) and mainly a defective peripheral tolerance checkpoint (in 7 out of 7 patients) have been demonstrated (Kinnunen et al., 2013a).

A critical question is whether tolerance defects can be reversed with targeted immunotherapy. In type 1 diabetes (T1D), treatment with the anti-CD20 monoclonal antibody (mAb) rituximab could not restore peripheral checkpoint function (Chamberlain et al., 2016). In contrast to continuous immunotherapy, immune reconstitution therapies (IRTs, alemtuzumab, cladribine, autologous hemopoietic stem cell transplantation) promote drug-free disease remission (Lünemann et al., 2020). The mAb alemtuzumab targets CD52+ immune cells (T and B cells, monocytes, natural killer (NK) cells, eosinophils and dendritic cells) causing their temporary depletion and renewal within weeks, months, or years, depending on the cell type. Clinically, alemtuzumab is very effective in diminishing MS disease activity, however its application is limited by frequent development of secondary autoimmune phenomena, most commonly thyroid-related (Coles et al., 2012).

CD52+ B cell depletion differs from CD20+ B cell depletion. First, CD52, but not CD20, is expressed on pre-B cells (Rodig et al., 2006), and accordingly anti-CD52 therapy affects a deeper depletion of B cells. Second, CD52—but not CD20 depletion—is followed by the generation of a high frequency of naïve B cells that exceeds baseline levels (Baker et al., 2017). To better understand efficacy and side effects of alemtuzumab, as well as B cell implication in RRMS and secondary autoimmunity, we evaluated the impact of alemtuzumab on B cell tolerance mechanisms in treated patients and controls. We first assessed the peripheral B cell tolerance checkpoint integrity with a validated method involving cloning of individual mature naïve B cells and examination of their poly- and autoreactivity. Next, we inquired whether next-generation sequencing (NGS)(Vander Heiden et al., 2017) of the BCR can identify alterations in tolerance mechanisms present in either immunotherapy-naïve RRMS patients or in patients treated with alemtuzumab. Finally, we examined if tolerance parameters correlate with clinical outcomes.

## 2. Materials and Methods

### 2.1. Patients, specimens, study protocol approval and patient consents

Peripheral blood was collected from healthy donors (HDs) and RRMS patients (immunotherapy-naïve or treated with alemtuzumab) after obtaining informed consent. Patients were diagnosed (Thompson et al., 2018), treated with alemtuzumab (12 mg/day iv for 5 consecutive days, followed one year later by 12 mg/day for 3 days), and followed at Eginitio University Hospital; blood samples were collected at least one year after the second treatment cycle. The three groups (HDs, immunotherapy-naïve RRMS and alemtuzumab-treated patients) were matched for age and sex. The study protocol and consents were approved by the Institutional Review Board of Eginitio Hospital. Two of the three healthy donor specimens (HD01 and HD02), were processed previously in the Laboratory of Dr. O’Connor at Yale University (Lee et al., 2016).

### 2.2. Immunophenotyping

PBMCs cryopreserved in liquid nitrogen vapors and 1mL aliquots of 10^7^cells/mL were thawed in 10 mL of 37°C RPMI medium with 10% fetal calf serum and let to rest for 30 minutes. 50 μL of PBMC-containing medium were stained with an antibody mix containing anti-human CD3-APC-AlexaFluor 750 (clone UCHT1), CD4-APC (13B8.2), CD8-FITC (B9.11), CD19-PC7(J3.119), CD16-ECD (3G8), CD25-APC-Alexa Fluor 700 (B1.49.9), CD127-PE (R34.34), CD45-Krome Orange(J.33) and 7AAD live/dead stain (all from Beckman-Coulter, Immunotech, USA), for 15 minutes at room temperature. For intracellular staining of FOXP3, cells were first stained with CD4-APC (13B8.2) and CD25-APC-Alexa Fluor 700 (B1.49.9) (Beckman-Coulter, Immunotech, USA), and then fixed, permeabilized and stained with anti-Hu-FOXP3-PE (F-9, Thermo Fischer Scientific, USA), according to the manufacturer’s instructions. Stained samples were analyzed on a 3-LASER, 13 color, DX Flex Flow Cytometer (Beckman-Coulter, USA).

### 2.3. PTPN22 R620W/C1858T variant genotyping

A previously described(Wardemann et al., 2003) protocol was applied. Briefly, DNA extraction from whole blood (Qiagen) was followed by PCR amplification of the R620W/C1858T variant-containing PTPN22 region (Thermo Fisher Scientific, Integrated DNA Technologies) and Sanger sequencing.

### 2.4. B cell isolation, staining and single-cell sorting

A previously established (Cotzomi et al., 2019; Wardemann et al., 2003) protocol was followed. Briefly, cryopreserved PBMCs were thawed, and B cells were positively selected using a biotin antibody cocktail with magnetic streptavidin nanobeads (MojoSort human pan B cell isolation kit, BioLegend). The isolated B cells were stained with anti-human CD10, CD19, CD27 (BioLegend), IgM, CD21, and IgG (BD Biosciences). Single mature naïve B cells (IgM^+^, IgG^−^, CD21^+^, CD27^−^, CD19^+^, CD10^−^) were then sorted into 96-well plates using a FACSAria flow cytometer and immediately frozen on dry ice and stored until further use.

### 2.5. mAb production

An established (Wardemann et al., 2003) approach was used. For each B cell, matching heavy (IgH) and light (IgK and IgL) chain variable immunoglobulin regions were amplified, cloned into expression vectors, sequenced, and analyzed with IgBlast (http://www.ncbi.nlm.nih.gov/projects/igblast). HEK 293A cells were then co-transfected with the heavy and light chain vectors, and the supernatant was applied to polyreactivity and, as purified IgG, autoreactivity ELISAs and the thyroid stimulating hormone receptor (TSHR) live cell-based assay (CBA). Each mAb was cloned, expressed and tested in duplicate.

### 2.6. ELISA

Established (Cotzomi et al., 2019; Wardemann et al., 2003) protocols were followed. Briefly, mAb concentrations were determined in HEK supernatants by sandwich ELISA using anti-human IgG antibodies (Jackson Laboratories). Supernatants were tested for polyreactivity, defined as reactivity against three different antigens: dsDNA, LPS, and insulin (all by Sigma-Aldrich). Autoreactivity was determined by application of purified mAbs to microplates precoated with human epithelial cell lysate (QUANTA Lite, INOVA Diagnostics).

### 2.7. Live CBA for the detection of Thyroid-Stimulating Hormone Receptor (TSHR) antibodies

A previously described CBA was used(Stathopoulos et al., 2019). In brief, HEK 293T cells were transfected with a plasmid vector encoding human TSHR-GFP (TSHR - Green Fluorescent Protein; kind gift from MABY and Dr. Bieberman of Charite University Hospital). Transfected cells were then incubated with 2 µg/mL of mAb, and subsequently with a secondary, Alexa fluor 647-labeled anti-human IgG antibody (1:1000, Jackson Laboratories). Fluorescence was measured with a BD FACS Canto cytometer. Positivity was assessed by calculating the delta mean fluorescence intensity (ΔMFI) index with FlowJo v10 (BD). The commercial anti-human TSHR mAb clone m22 (ABCDY antibodies, kind gift from MABY) served as a positive control. The cutoff for positivity was applied at the control mean + 4SDs.

### 2.8. cDNA libraries construction and BCR sequencing

RNA was isolated from PBMCs with the QIAwave RNA Mini Kit (Qiagen). cDNA libraries were prepared with the New England Biolabs Human Next^®^ Immune Sequencing Kit(Vander Heiden et al., 2017). Reverse transcription employed biotin-labeled primers and addition of a unique molecular identifier (UMI). cDNA was purified using streptavidin magnetic beads, and IgH and IgK/IgL or just IgH variable regions were amplified by a series of two PCRs. DNA libraries were pooled equimolarly, followed by high throughput paired-end sequencing on either the Illumina MiSeq or NextSeq (Functional Genomics Center Zurich, ETH and University of Zurich).

### 2.9. Raw data quality control

Quality control was performed using a slightly modified pRESTO workflow (Vander Heiden et al., 2017). First, reads with a mean Phred quality score >20 and without a valid C-region primer were discarded. Both IgH and IgK/IgL primers were used to improve sampling and increase the number of analyzed reads. Reads with identical UMIs were categorized into a single consensus sequence. If multiple isotypes were identified within a single UMI read group, the consensus sequence was generated using only the reads assigned to the majority isotype. UMI consensus sequence mate-pairs were initially assembled de novo into full-length Ig sequences, requiring a minimum overlap length of 8 bp, a maximum error rate of 0.3, and a p-value threshold of 1 × 10⁻□. Mate-pairs that could not be assembled de novo were instead aligned to the 202248-3 release of the International Immunogenetics Information System (IMGT) V-segment germlines, with a minimum identity threshold of 0.5 and an E-value cutoff of 1 × 10⁻□. A secondary isotype assignment was performed by aligning the tail of the UMI consensus sequences to a set of sequences anticipated to be upstream of the C region primer. These reference sequences were derived from the IMGT C region and J segment consensus database. Following preprocessing, V(D)J germline segments were assigned, using the IgBLAST search algorithm (Ye et al., 2013).

### 2.10. BCR repertoire analysis

Reported germline sequences, combined with the C-region annotation, were used to define the locus and isotype. IgBLAST results were parsed and standardized using ChangeO (Vander Heiden et al., 2017), and filtered to include only productive BCR sequences. Genotyping was performed using TIgGER (Vander Heiden et al., 2017). Germline sequences of each assigned clonal group were identified using Change-O. The genotyped, clonal-assigned reads were used to calculate somatic hypermutations (SHMs) with SHazaM, by comparing observed sequences against the germline. With the SHM frequencies calculated, a 1% (of nucleotides) mutational cutoff was set to separate naive and memory BCRs (for both IgH and IgK/IgL). V- and J-gene (IgH and IgK/IgL) family usage was calculated as the percentage of unique sequences assigned to a given gene family within the total number of sequences for each isotype in each compartment (naïve/memory). Variations in the usage of IgK V-gene sequences were evaluated by calculating the proportion of upstream (distal) vs. downstream (proximal) V-gene families. The definition of IgK V-gene sequences as upstream or downstream was based on annotations provided by IMGT. IgH CDR3 properties (Vander Heiden et al., 2017; Wu et al., 2010) (amino acid sequence length, hydrophobicity -estimated as the grand average hydropathy index, or GRAVY-, net charge, polarity, acid chain bulkiness, aliphatic index, as well as the ratios of aromatic, acidic, and basic amino acid residues) were measured separately for naive and memory sequences using Alakazam (Vander Heiden et al., 2017).

### 2.11. Statistical analysis

Data were plotted and analyzed with the GraphPad Prism software version 8.3.1. Unpaired t-tests and one-way ANOVA tests were performed to compare means between different groups.

## 3. Results

### 3.1. Study subjects, immunophenotyping, and production of mAbs

Study subjects included five healthy donors, four immunotherapy-naive RRMS patients, and nine alemtuzumab-treated RRMS patients (**Table**). Age and sex distribution did not differ significantly between the three patient groups. RRMS patients were sampled after an attack leading to diagnosis but prior to steroids and all tested positive for intrathecal immunoglobulin synthesis. Alemtuzumab-treated patients were sampled 3.5 to 4.8 years [mean ± standard deviation (SD) 3.8 ± 0.39 years] post treatment initiation. Moreover, immunophenotyping at the time of sampling did not reveal striking differences in immune reconstitution between alemtuzumab-treated patients (**Supplementary Table 2**). At the time of sampling, six of the alemtuzumab-treated patients had suffered secondary autoimmune complications, while two patients showed evidence of MS activity post-treatment (**Table**). Four patients, free of MS activity at the time of sampling, developed such activity at a later time point (**Supplementary Table 1**). To assess B cell tolerance mechanisms, we produced 138 mAbs in biological duplicates from single mature naïve B cells (IgM^+^, IgG^−^, CD21^+^, CD27^−^, CD19^+^, CD10^−^) of 12 subjects. Sequences of the cloned variable regions were compared to the germline using IgBLAST, and mutated sequences were excluded from the analysis as previously described (Cotzomi et al., 2019) (**Supplementary Table 3**). Importantly, carriers of the C1858T PTPN22 polymorphism, associated with defective central and peripheral B cell tolerance checkpoint function (Kinnunen et al., 2013a), were excluded from all subject groups.

**Table.** Characteristics of study subjects

| **Samples** | **Age range<sup>1</sup>** | **Sex** | **Time to sampling<sup>2</sup>** | **MS activity** | **Secondary autoimmunity** | **Tolerance assay** | **Mature naïve autoreactive fraction (vs. HDs)** | **BCR NGS** |
| --- | --- | --- | --- | --- | --- | --- | --- | --- |
| **HD01<sup>3</sup>** | 26-30 | F | NA | NA | NA | Yes | NA | No |
| **HD02<sup>3</sup>** | 46-50 | M | NA | NA | NA | Yes | NA | No |
| **HD13** | 31-35 | F | NA | NA | NA | No | NA | Yes |
| **HD18** | 36-40 | F | NA | NA | NA | Yes | NA | Yes |
| **HD19** | 41-45 | M | NA | NA | NA | No | NA | Yes |
| **RRMS09** | 46-50 | F | NA | NA | NA | No | NA | Yes |
| **RRMS16** | 41-45 | F | NA | NA | NA | Yes | High | Yes |
| **RRMS17** | 26-30 | F | NA | NA | NA | Yes | High | Yes |
| **RRMS21** | 26-30 | M | NA | NA | NA | Yes | Same | Yes |
| **Ale27** | 36-40 | F | 3.9 | No | No | Yes | Low | Yes |
| **Ale40** | 36-40 | F | 3.6 | No | No | Yes | High | Yes |
| **Ale46** | 31-35 | F | 3.9 | No | No | Yes | Same | No |
| **Ale03** | 36-40 | M | 3.9 | No | Yes (Graves) | Yes | High | Yes |
| **Ale25** | 26-30 | F | 3.7 | No | Yes (Graves) | Yes | Low | Yes |
| **Ale26** | 51-55 | F | 3.8 | No | Yes (Hashimoto) | No | NA | Yes |
| **Ale33** | 36-40 | M | 3.5 | No | Yes (Hashimoto) | Yes | High | No |
| **Ale24** | 41-45 | M | 3.5 | Yes | Yes (ITP, neutropenia, Hashimoto) | No | NA | Yes |
| **Ale29** | 36-40 | M | 4.8 | Yes | Yes (ITP) | No | NA | Yes |

<sup>1</sup>Age range at the time of sample acquisition. <sup>2</sup>Time to sampling in years was defined as the interval between initial dose of alemtuzumab and collection of the sample. <sup>3</sup>Studied by the laboratory of KC O'Connor at Yale University (Lee et al., 2016). None of the subjects carried the PTPN22 R620W polymorphism. Ale: Alemtuzumab-treated RRMS patients; F: Female; HD: Healthy donor; ITP: Immune Thrombocytopenia; M: Male; NA: Not applicable; RRMS: Relapsing-remitting multiple sclerosis.

### 3.2. Polyreactive fractions of mature naïve B cells from HDs, RRMS patients, and alemtuzumab-treated RRMS patients

To determine whether anti-CD52 treatment with alemtuzumab affects the frequency of polyreactive mature naïve B cells, we used an established and extensively validated experimental approach that tests reactivity of mature naïve B cell-derived mAbs against three antigens: dsDNA, LPS, and insulin, with polyreactivity being defined as reactivity against all three antigens. The fraction of polyreactive mAbs ranged from 9% to 14.29% in HDs (mean ± SD 10.57 ± 3.24%), from 7.29% to 12.50% in immunotherapy-naïve RRMS patients (10.9 ± 2.7%), and from 8.33% to 33.33% in patients with RRMS treated with alemtuzumab (15.31 ± 9.2%) (**Fig. 1**). Polyreactive mAb percentages did not differ significantly between the three groups.

**Figure 1.**
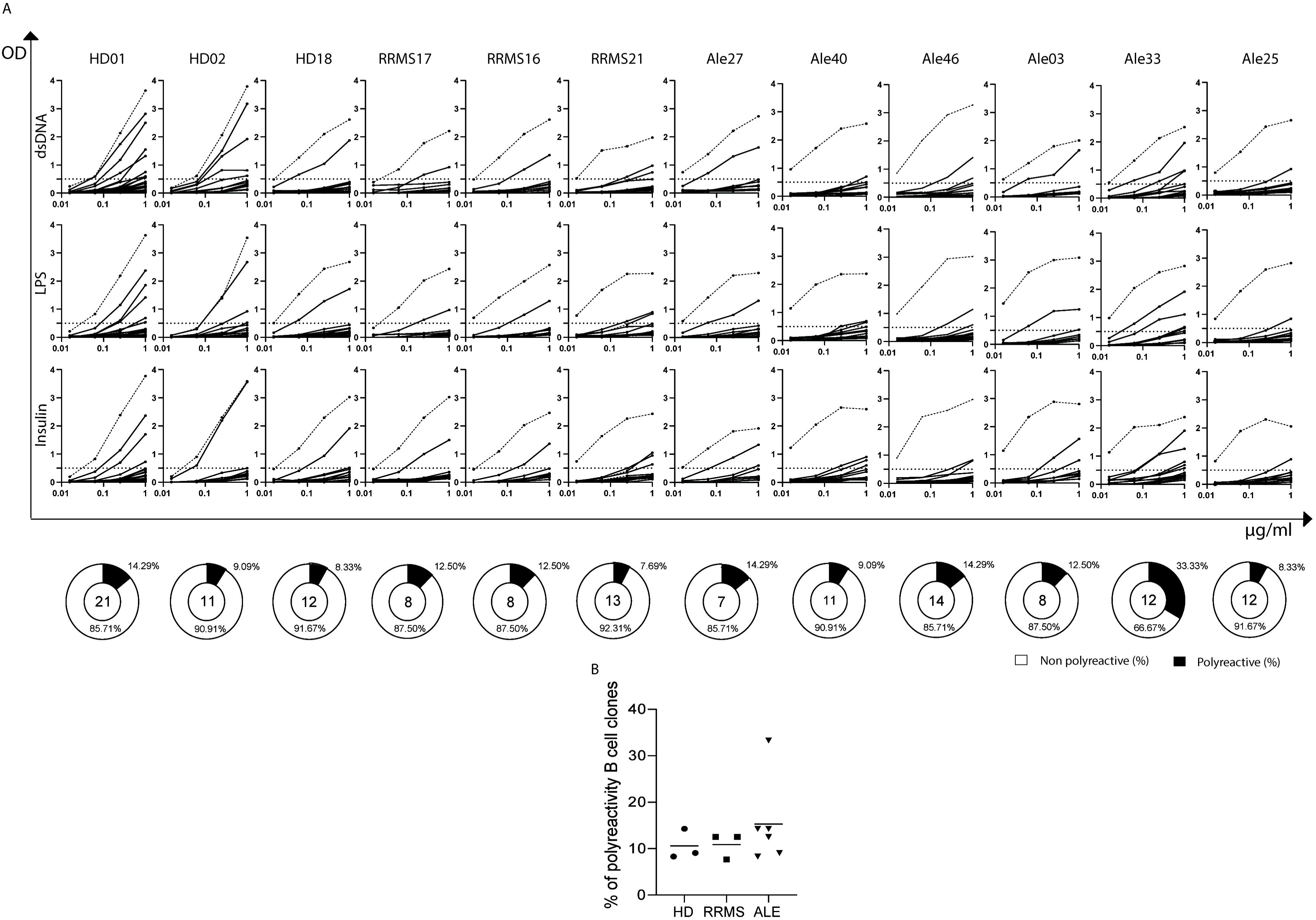
Mature naïve B cell polyreactivity. **(A)** Mature naïve B cell-derived mAbs from 3 HDs, 3 immunotherapy-naïve and 6 alemtuzumab-treated RRMS patients were tested for their reactivity against dsDNA, LPS, and insulin by ELISA. ODs are shown on the y axis and mAbs concentrations on the x axis. Solid lines connect OD values of each mAb at a given dilution, and each point represents the mean of biological duplicates. Dotted curves represent positive control mAb ED38 binding; horizontal dotted lines represent the positivity cutoff. Below, black-filled areas of pie charts indicate the frequency of polyreactive mAbs for each subject, while the number in the center of each pie chart indicates the number of cloned mAbs per subject. (**B)** Polyreactivity frequencies in HDs, immunotherapy-naïve and alemtuzumab-treated RRMS patents, with lines representing mean values. Means did not differ significantly. Ale: Alemtuzumab-treated RRMS patients; dsDNA: Double-stranded DNA; HD: Healthy donor; LPS: Lipopolysaccharide; mAb: monoclonal antibody; RRMS: Relapsing-remitting multiple sclerosis.

### 3.3. Autoreactive fractions of mature naïve B cells from HDs, RRMS patients, and alemtuzumab-treated RRMS patients

The peripheral B cell tolerance checkpoint integrity was assessed by examining purified, mature naïve B cell-derived mAbs for potential autoreactivity, i.e. reactivity against human epithelial type two (HEp-2) cell lysate (Kinnunen et al., 2013a) (**Fig. 2**). After performing a historically validated HEp-2 ELISA, we found that the percentage of autoreactive mAbs ranged from 19.05 to 33.33% in HDs (mean ± SD 26.55 ± 7.17%), from 23.08 to 37.5% (32.7 ± 8.33%) in immunotherapy-naïve RRMS patients, and from 8.33 to 58.33% (30.56 ± 17.98%) in alemtuzumab-treated patients. Autoreactive mAb percentages did not differ significantly between the three groups.

**Figure 2.**
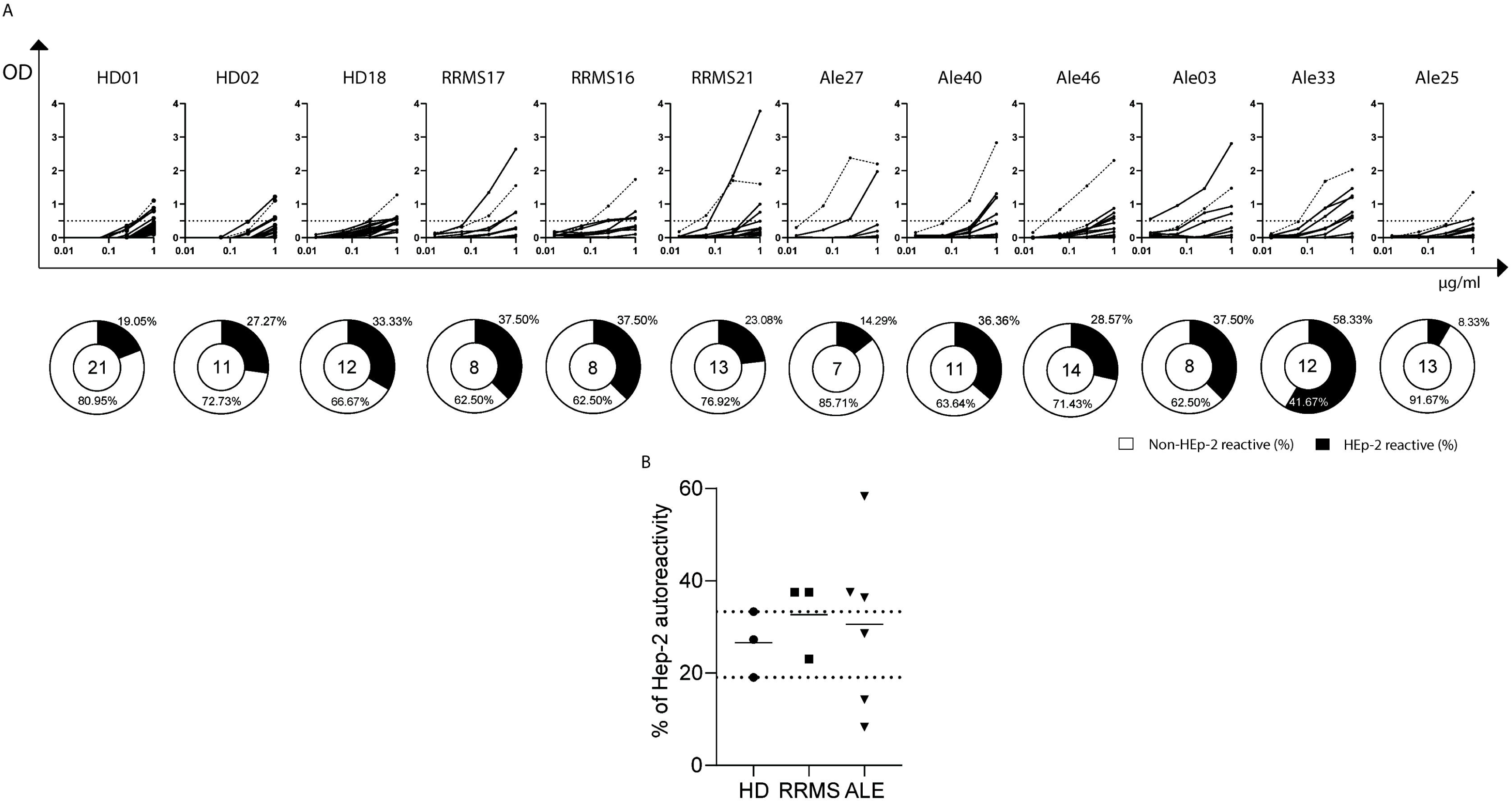
Mature naïve B cell autoreactivity. **(A)** Mature naïve B cell-derived mAbs from 3 HDs, 3 immunotherapy-naïve and 6 alemtuzumab-treated RRMS patients were tested for their reactivity against HEp-2 cell lysate. ODs are shown on the y axis and mAbs concentrations on the x axis. Solid lines connect OD values of each mAb at a given dilution, and each point represents the mean of biological duplicates. Dotted curves represent positive control mAb ED38 binding; horizontal dotted lines represent the positivity cutoff. Below, black-filled areas of pie charts indicate the frequency of autoreactive mAbs for each subject, while the number in the center of each pie chart indicates the number of cloned mAbs per subject. **(B)** Autoreactivity frequencies in HDs, immunotherapy-naïve and alemtuzumab-treated RRMS patents, with lines representing mean values. Means did not differ significantly. Ale: Alemtuzumab-treated RRMS patients; HEp-2: Human Epithelial Type 2 cells; HD: Healthy donor; RRMS: Relapsing-remitting multiple sclerosis.

Previously, autoreactivity of mature naïve B cells was found to be significantly increased in RRMS compared to HDs, and no overlap was observed between HD and RRMS mature naïve autoreactive fractions, although separation was borderline (HD <30% and RRMS 33.3 to 45.5%; n=11 HDs and 7 RRMS patients)(Kinnunen et al., 2013a; Meffre and O’Connor, 2019). Here, some overlap did occur and HD autoreactivity ranged from 19.05 to 33.33% while RRMS autoreactivity from 23.08 to 37.5%. Moreover, determination of checkpoint integrity is often dichotomous (preserved/damaged) and made on an individual subject basis (Chamberlain et al., 2016; Cotzomi et al., 2019; Glauzy et al., 2017; Kinnunen et al., 2013a; Lee et al., 2016; Samuels et al., 2005; Yurasov et al., 2005). In that regard, the function of the peripheral checkpoint was likely compromised in 2 of 3 RRMS patients (**Fig. 2**), in line with previous results of a compromised peripheral checkpoint in 7 out of 7 patients.

In alemtuzumab-treated patients, three of six demonstrated a percentage of autoreactive naïve B cells higher than that of HDs (36.36% in Ale40, 37.50% in Ale03, and 58.33% in Ale33), likely pointing to defective peripheral B cell tolerance function (**Fig. 2**). In contrast, two of six alemtuzumab-treated patients demonstrated a fraction of autoreactive naïve cells lower than that of HDs (14.29% in Ale27 and 8.33% in Ale25). This variability did not align with secondary autoimmunity and future MS activity (**Table 1** and **Supplementary Table 1**, respectively).

### 3.4. Mature naïve-derived mAbs from RRMS patients treated with alemtuzumab do not bind Graves’ disease autoantigen TSHR

Almost 50% of alemtuzumab-treated RRMS patients develop secondary, thyroid-related autoimmunity, commonly TSHR autoantibody-positive Graves’ disease (Coles et al., 2012). With that in mind, we asked whether mature naïve B cell-derived mAbs from the two RRMS patients treated with alemtuzumab that developed TSHR autoantibody-positive Graves’ disease bound TSHR. Of note, only one of these two patients had a high fraction of autoreactive mature naïve B cells. mAbs produced from mature naïve B cells of HDs, immunotherapy-naïve RRMS patients and alemtuzumab-treated patients that did not develop Graves’ disease served as negative controls. A commercial mAb against the TSHR was used as a positive control (Sanders et al., 2003) (**Supplementary Fig. 1**). None of the 95 mAb tested bound the TSHR (**Supplementary Table 4**). indicating that even in the one patient with a high percentage of autoreactive mature naïve B cells, autoimmunity likely initiates beyond the lymphnode/T cell help stage of B cell development and not in the naïve stage, prior to the encounter of the (auto)antigen.

### 3.5. Exploratory analysis of naïve BCR repertoire properties as markers of tolerance

As the established approach to assess B cell tolerance is time-consuming and laborious, we sought to investigate whether BCR NGS could evaluate B cell tolerance checkpoint integrity to a similar extent or perhaps with enhanced sensitivity. To that end, we performed BCR NGS out of total PBMC RNA for the indicated HD, immunotherapy-naïve RRMS and alemtuzumab-treated RRMS samples (**Table**). After raw data quality control, we applied a 1% mutational cutoff to separate naïve from memory IgH sequences, yielding percentages comparable(Morbach et al., 2010) to immunophenotypic profiling of HDs (**Supplementary Table 5**). With that separation, we were able to reproduce known differences in CDR3 properties between memory and naïve B cells (Wu et al., 2010) (**Supplementary Fig. 2**).

Several reports (Bridges et al., 1995; Samuels et al., 2005; Shiokawa et al., 1999; Wardemann et al., 2003; Wu et al., 2010) have indicated that IgH CDR properties might be a measure for autoreactivity and compromised B cell tolerance; therefore we examined whether these differ between in the naïve compartment of HDs, immunotherapy-naïve RRMS patients and RRMS patients treated with alemtuzumab (**Fig. 3A-D**). Of the four CDR3 properties examined, one was found to differ significantly between subgroups. Specifically, net charge was found to be lower in alemtuzumab-treated RRMS patients compared to HDs (IgD+IgM+ mean ± SD −0.49 ± 0.04 vs. −0.38 ± 0.036, P = 0.0036). We examined if CDR3 properties correlate with high naïve B cell autoreactivity as determined by ELISA or MS activity/secondary autoimmunity up to the sampling timepoint; this however was not the case (**Supplementary Fig. 3, 4)**

**Figure 3.**
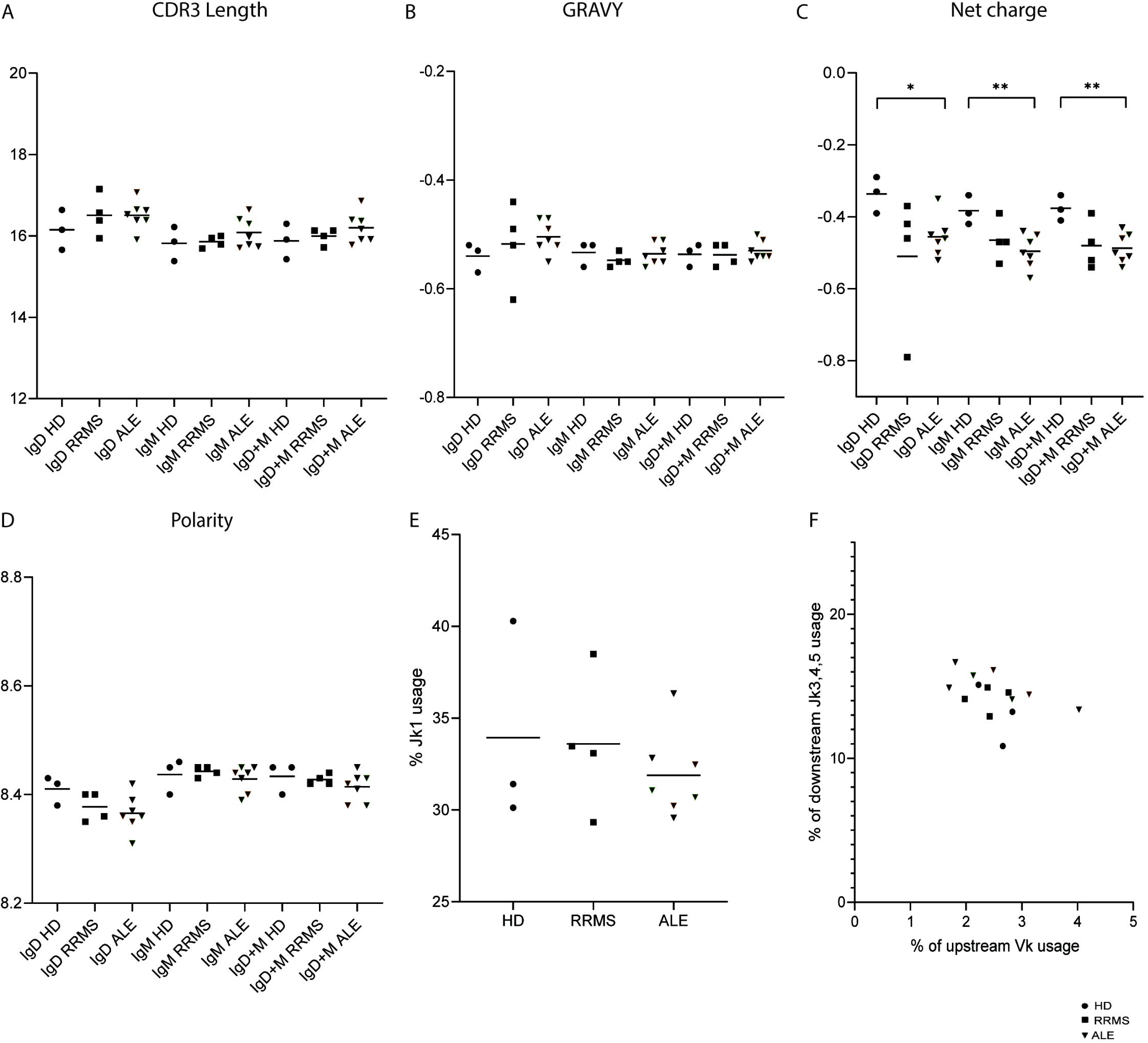
Next generation sequencing analysis of naïve B cell receptor repertoire. **(A-D)** IgH CDR3 properties, calculated per Ig class: **(A)** CDR3 length, **(B)** the grand average hydropathy index (GRAVY), **(C)** net charge, **(D)** polarity. Black circles represent individual HD values, black squares those of immunotherapy-naïve RRMS patients and black triangles those of alemtuzumab-treated RRMS patients. **(E-F)** Usage of Jk1, upstream Vk, and downstream Jk3,4,5 gene segments of Igk: **(E)** collective data on Jk1 gene usage frequency across all study groups, **(F)** upstream Vk gene usage frequency plotted against downstream Jk3,4,5 usage in naive B cells from all study subjects. Differences between groups (HD, immunotherapy-naïve RRMS, and alemtuzumab-treated RRMS) were tested with the Mann-Whitney test, and p values are indicated if significant (**P ≤ 0.01; *P ≤ 0.05). ALE: Alemtuzumab-treated RRMS patients; CDR: Complementarity-Determining Region; HD: Healthy donor; Ig: immunoglobulin; Igk: kappa light chain; IgH: heavy chain; NGS: Next-generation sequencing; RRMS: Relapsing-remitting multiple sclerosis.

Next, we examined whether signs of altered secondary VJk recombination exist. Of note, VJ recombination in the Igk locus occurs in two stages: the primary, generating diversity; and the secondary, replacing autoreactive BCRs with non-reactive ones in the context of B cell tolerance mechanisms (Nemazee, 2017). Increased repertoire frequency of the Jk1 gene segment has been associated with increased frequency of primary recombination, while increased frequency of the downstream Jk3,4,5 gene segments, as well as of the upstream Vk genes with increased secondary rearrangements (Mensah et al., 2019; Nemazee, 2017). Here, Jk1 frequency did not significantly differ between HDs, immunotherapy-naïve, and alemtuzumab-treated RRMS patients (**Fig. 3E**) while naïve B cell usage of upstream Vk and downstream Jk3,4,5 gene segments did not consistently separate subject groups (**Fig. 3F**).

### 3.6. Exploratory analysis of memory BCR repertoire properties

As a third B cell tolerance checkpoint might exist between the naïve and the memory IgM compartment (Tsuiji et al., 2006), we examined memory B cell repertoire characteristics as well. We first analyzed CDR3 properties in the three subject groups (HDs, immunotherapy-naïve and alemtuzumab-treated RRMS) per immunoglobulin class (**Fig. 4**). Significant differences were observed in the IgD compartment. Specifically, alemtuzumab-treated RRMS patients showed a significantly increased CDR3 length (mean ± SD 16.18 ± 0.6) compared to HDs (15.07 ± 0.24; P = 0.012). Moreover, polarity was significantly decreased in alemtuzumab-treated patients (8.4 ± 0.1) compared to both HDs (8.6 ± 0.06; P = 0.039) and RRMS patients (8.5 ± 0.06; P = 0.045) (**Fig. 4A-D**). Taken together, these differences represent interesting findings of an exploratory analysis that warrant confirmation in larger studies.

**Figure 4.**
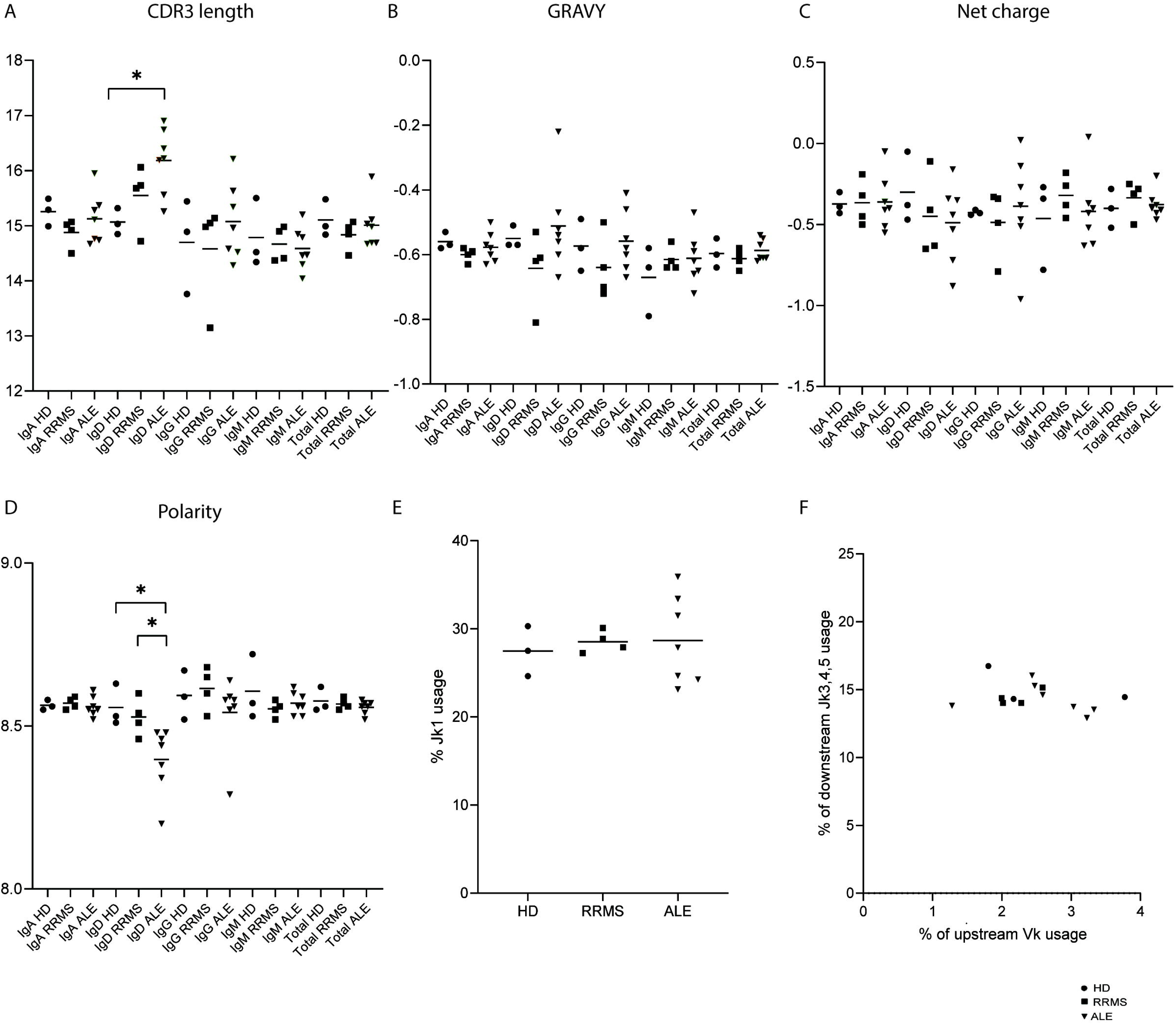
Next generation sequencing analysis of memory B cell receptor repertoire. IgH CDR3 properties, calculated per Ig class: **(A)** CDR3 length, **(B)** the grand average hydropathy index (GRAVY), **(C)** net charge, **(D)** polarity. Black circles represent individual HD values, black squares those of immunotherapy-naïve RRMS patients and black triangles those of alemtuzumab-treated RRMS patients. **(E-F)** Usage of Jk1, upstream Vk, and downstream Jk3,4,5 gene segments of Igk: **(E)** collective data on Jk1 gene usage frequency across all study groups, **(F)** upstream Vk gene usage frequency plotted against downstream Jk3,4,5 usage in memory B cells from all study subjects. Differences between groups (HD, immunotherapy-naïve RRMS, and alemtuzumab-treated RRMS) were tested with the Mann-Whitney test, and p values are indicated if significant (**P ≤ 0.01; *P ≤ 0.05). ALE: Alemtuzumab-treated RRMS patients; CDR: Complementarity-Determining Region; HD: Healthy donor; Ig: immunoglobulin; Igk: kappa light chain; IgH: heavy chain; NGS: Next-generation sequencing; RRMS: Relapsing-remitting multiple sclerosis.

Similar to the naïve compartment, Jk1 frequency did not differ between HDs, immunotherapy-naïve, and alemtuzumab-treated RRMS patients (**Fig. 4E**). Further, naïve B cell usage of upstream Vk and downstream Jk3,4,5 gene segments did not consistently separate subject groups (**Fig. 4F**).

## 4. Discussion

We examined B cell tolerance mechanisms in the setting of RRMS and in vivo perturbation by alemtuzumab; the effect of transient CD52+ immune cell depletion on tolerance checkpoints had not been researched before. In addition, we combined an approach used historically for analysis of B cell checkpoint function involving construction of recombinant mAbs from post-checkpoint B cells with repertoire analysis by NGS. Again, this combination had not been applied —to our knowledge— beforehand. Of particular importance, our results may be relevant not only to immune reconstitution with alemtuzumab, but also to other immune reconstitution therapies such as cladribine and B cell-targeted therapies (Lünemann et al., 2020).

Although we examined both poly-and autoreactivity of mature naïve B cells, it is autoreactivity that better defines peripheral B cell tolerance checkpoint function (Wardemann et al., 2003). After performing autoreactivity ELISA with mature naïve-derived mAbs, we did not find significant differences in mean autoreactive fractions between the three subgroups (HDs, immunotherapy-naïve and alemtuzumab-treated RRMS), pointing to absence of a major effect of transient CD52 depletion with alemtuzumab on peripheral B cell checkpoint function. Dichotomous and individual examination of peripheral B cell tolerance checkpoint integrity pointed to compromised checkpoint function in 2 out of 3 immunotherapy-naïve RRMS patients, in line with previous results (Kinnunen et al., 2013a; Meffre and O’Connor, 2019), and In, 3 out of 6 alemtuzumab-treated patients. Two alemtuzumab-treated patients showed particularly low autoreactive fractions, lower than those of all HDs and immunotherapy-naïve RRMS patients. High or low autoreactivity of alemtuzumab-treated patients however did not correlate with secondary autoimmunity or future MS activity and appeared to be a random, stochastic phenomenon associated with immune reconstitution.

In some similarity, T1D patients treated with the anti-CD20 mAb rituximab did not regain function of the defective peripheral B cell tolerance checkpoint (Chamberlain et al., 2016). Although diseases and therapies differ, similarities include an autoimmune setting and therapeutic immune reconstitution. Compared to rituximab, alemtuzumab induces deeper depletion of B cells and more effectively depletes T cells including Tregs, which are known to influence peripheral checkpoint function (Janssen et al., 2014; Kinnunen et al., 2013b; Meffre and O’Connor, 2019). In addition, Treg function is compromised in MS but restored post alemtuzumab application (Viglietta et al., 2004; Dominguez-Villar et al., 2011; Haas et al., 2019).

We additionally investigated whether defects in peripheral tolerance can directly lead to secondary autoimmunity, which often targets the thyroid in the form of Graves’ disease and anti-TSHR antibodies (Coles et al., 2012). We therefore tested mature naïve-derived mAbs against TSHR by CBA, however none of the tested mAbs showed binding with TSHR. Although we did not find evidence that TSHR autoantibody generation is a pre-germinal center event, this cannot be excluded since autoantigen-specific B cells are rare.

The IgH CDR3 region plays a critical role in determining specificity toward antigens as well as repertoire diversity. Therefore, its composition and distinct physicochemical properties are of interest in the study of autoimmunity and may correlate with B cell tolerance defects (Lee et al., 2016; Stathopoulos et al., 2018; Vander Heiden et al., 2017; Xu and Davis, 2000). Previous studies examining autoimmune B cell repertoires have shown that IgH CDR3 regions of patients are longer (Larimore et al., 2012; Wardemann et al., 2003), more hydrophobic (Meffre et al., 2004), and more positively charged (Richardson et al., 2013) compared to controls. Here, we found that in the naïve compartment of alemtuzumab-treated RRMS patients, CDR3 net charge is significantly lower than in HDs, an effect that can be attributed to RRMS, alemtuzumab treatment, or both. Of interest and potential biological significance was also the fact that while CDR3s were of shorter length in HD memory (primarily IgM) vs. HD naïve cells as previously reported (Wu et al., 2010)(**Supplementary Fig. 2**), they tended to be longer in the RRMS vs. HD naïve compartment (**Fig. 3**). As IgM memory B cells and plasma cells tend to be less poly- and autoreactive (Scheid et al., 2011; Tsuiji et al., 2006) (perhaps as a result of affinity maturation and selection pressure), longer naïve CDR3s in RRMS may represent a sensitive marker of promiscuity/polyreactivity. Indeed, such antibody/B cell promiscuity has been reported previously (Brändle et al., 2016; Quintana et al., 2012, 2008; Willis et al., 2015) in RRMS mAbs, serum and CSF.

Because memory cells very likely play a critical role in MS immunology (Jelcic et al., 2018) and as a third B cell tolerance checkpoint might exist between the naïve and the memory IgM compartment (Tsuiji et al., 2006), we also examined whether memory IgH CDR3 properties differ between HDs and immunotherapy-naïve or alemtuzumab-treated RRMS patients. Indeed, we found significant differences in IgD CDR3 length, and polarity between alemtuzumab-treated RRMS patients and HDs. Specifically, both RRMS and more so alemtuzumab treatment were associated with longer CDR3s. Interestingly, a similar effect was observed in RA patients treated with rituximab, where treatment was associated with longer IgD memory CDR3s and a lower IgD memory mutation acquisition rate (Muhammad et al., 2009). Therefore, the increase in CDR3 length might have different etiologies in RRMS and in alemtuzumab-treated RRMS. In the latter case of alemtuzumab −similarly to the case of rituximab−, it likely is the transient B cell depletion leads to a repopulation similar to ontogeny, known to be associated with longer CDR3s (Souto-Carneiro et al., 2005).

Overall, we conducted a study whose main strength is the simultaneous, in-depth analysis of B cell reactivity and BCR repertoire diversity in an in vivo perturbed setting of drug-induced immune reconstitution. Simultaneous measurement of both parameters in the same subjects allowed for examination of their correlation. Moreover, our study of disease and treatment mechanisms was performed with human samples and therefore our results illuminate directly both RRMS immunopathology as well as CD52-mediated immune reconstitution. In addition, our findings may offer insight to immune reconstitution approaches beyond alemtuzumab such as cladribine and B cell-depleting therapies. Weaknesses include lack of longitudinal pre- and post-treatment samples, as alemtuzumab’s application in our center was stopped due to a less favorable risk/benefit ratio compared to other treatments of RRMS such as B cell depletion. We compensated this lack with thorough matching cases with controls in regard to disease activity, sample acquisition time point, sex, and age. Finally, our BCR NGS experiments are of an exploratory nature and warrant confirmation in larger studies.

Our investigations show for the first time that transient CD52+ immune cell depletion does not affect major changes of peripheral B cell tolerance checkpoint function in RRMS patients. Mature naive autoreactivity of alemtuzumab-treated patients varied widely, but this variability did not relate to presence of secondary autoimmunity, future MS activity or Treg frequency and therefore most likely represents a stochastic phenomenon. Secondary autoimmunity targeting the thyroid (up to the time of sample acquisition) did not correlate with any of the measured B cell parameters and likely is a post-germinal center effect as conspicuous memory B cell expansions co-appear with autoimmune events (Ruck et al., 2022).

## Abbreviations

AIRE: Autoimmune Regulator
BCR: B Cell Receptor
CBA: Cell-Based Assay
CDR3: Complementarity-Determining Region 3
GFP: Green Fluorescent Protein
HDs: Healthy Donors
Hep-2: Human Epithelial Cells Type 2
IgH: Immunoglobulin Heavy Chain
IgK: Immunoglobulin Kappa Light Chain
IgL: Immunoglobulin Lambda Light Chain,
IRTs: Immune Reconstitution Therapies
MS: Multiple Sclerosis
mAb: Monoclonal Antibody
NGS: Next-Generation Sequencing
PBMCs: Peripheral Blood Mononuclear Cells
RRMS: Relapsing-Remitting Multiple Sclerosis
SHMs: Somatic Hypermutations
SLE: Systemic Lupus Erythematosus
T1D: Type 1 Diabetes
Tregs: Regulatory T Cells
TSHR: Thyroid-Stimulating Hormone Receptor

## Data availability

Anonymized data will be shared to any qualified investigator by the corresponding author upon reasonable request.

## Acknowledgments

The authors thank NEB for providing a Next® Immune Sequencing Kit, Chen Song of NEB for her valuable assistance with next generation sequencing quality control, Dr. H. Rehrauer of the Functional Genomics Center Zurich of the ETH and the University of Zurich for his assistance with the physical aspect of next generation sequencing, Drs. Steven Kleinstein and Gisela Gabernet of Yale University for valuable discussions, Dr. Sofia Grammenoudi from the BSRC Fleming cell sorting core facility for her support, and Dr. A. Goules from the Department of Pathophysiology of the National and Kapodistrian University of Athens for providing bacterial culture resources. The graphical abstract was created with Biorender.

