## Supplementary for "B cell tolerance checkpoint function in multiple sclerosis and transient CD52 depletion"

**Supplementary material**

**Contents**

**Supplementary Table 1.** Follow-up of alemtuzumab-treated patients post sample acquisition.

**Supplementary Table 2.** Immunophenotyping of alemtuzumab-treated patients at the time of monoclonal antibody production.

**Supplementary Table 3.** Sequence and reactivity tables for HDs, RRMS, and alemtuzumab-treated (ALE) specimens.

**Supplementary Table 4.** TSHR cell-based assay (CBA) results for HD, RRMS, alemtuzumab-treated RRMS patients.

**Supplementary Table 5.** BCR sorting into naive and memory sequences based on mutation counts for samples.

**Supplementary Figure 1.** Representative TSHR cell-based assay (CBA) flow cytometry plots.

**Supplementary Figure 2.** IgH CDR3 property comparisons between naïve and memory B cells of HDs.

**Supplementary Figure 3.** Analysis of naïve IgH CDR3 properties highlighting patients with a high fraction of autoreactive naïve B cells.

**Supplementary Figure 4.** Analysis of naïve IgH CDR3 properties highlighting MS activity/secondary autoimmunity up to the timepoint of sampling.

**Supplementary Table 1.** Follow-up of alemtuzumab-treated patients post sample acquisition

| **Patient** | **Alemtuzumab 1^st^ dose** | **Sample drawn** | **t from Ale to sample (y)** | **MS activity [t of sample]** | **Secondary autoimmunity [t of sample]** | **Last FU** | **MS activity [t of last FU]** | **Secondary autoimmunity [t of last FU]** | **Mature naïve autoreactive fraction (vs. HDs)** |
| --- | --- | --- | --- | --- | --- | --- | --- | --- | --- |
| Ale27 | 18.12.17 | 01.11.21 | 3.9 | No | No | 21.01.24 | Yes | No | Low |
| Ale40 | 06.05.19 | 13.12.22 | 3.6 | No | No | 24.01.23 | Yes | No | High |
| Ale46 | 15.07.19 | 01.06.23 | 3.9 | No | No | 08.07.25 | Yes | No | Same |
| Ale03 | 27.03.17 | 02.02.21 | 3.9 | No | Graves | 08.04.24 | Yes | Graves | High |
| Ale25 | 12.02.18 | 25.10.21 | 3.7 | No | Graves | 23.01.25 | No | Graves | Low |
| Ale26 | 29.01.18 | 01.11.21 | 3.8 | No | Hashimoto | 04.04.24 | No | Hashimoto | NA |
| Ale33 | 17.12.18 | 30.06.22 | 3.5 | No | Hashimoto | 25.04.25 | No | Hashimoto | High |
| Ale24 | 16.04.18 | 15.10.21 | 3.5 | Yes | ITP, neutropenia, Hashimoto | 08.10.24 | Yes | ITP, neutropenia, Hashimoto | NA |
| Ale29 | 31.07.17 | 01.07.22 | 4.8 | Yes | ITP | 03.02.25 | Yes | ITP | NA |
| Ale: alemtuzumab; t: time; FU: follow-up; ITP: immune thrombocytopenia. | | | | | | | | |  |

**Supplementary Table 2.** Immunophenotyping of alemtuzumab-treated patients at the time of monoclonal antibody production.

|  | **Ale27** | **Ale40** | **Ale46** | **Ale03** | **Ale33** | **Ale25** |
| --- | --- | --- | --- | --- | --- | --- |
| Viable cells (%of CD45+) | 91.66 | 77.09 | 97.66 | 96.56 | 96.3 | 96.24 |
| CD3+CD4+ (% of viable cells) | 49 | 46.4 | 48.7 | 40.8 | 42.9 | 35.5 |
| CD3+CD8+(% of viable cells) | 15.33 | 18 | 21.92 | 24.5 | 25.65 | 23.78 |
| CD19+(% of viable cells) | 11.53 | 19.86 | 14.6 | 23.03 | 15.28 | 23.46 |
| CD3-CD16+(% of viable cells) | 20.46 | 10.58 | 9.46 | 4.91 | 11.13 | 11.81 |
| CD25+CD127-(% of viable cells) | 5.49 | 6.15 | 3.94 | 6.22 | 4.39 | 4.65 |
| FOXP3+ (% of viable CD4+ cells) | 2.96 | 3.65 | 2.54 | 1.6 | 1.75 | 3.35 |

**Supplementary Table 3.** Sequence and reactivity tables for HDs, immunotherapy-naïve RRMS patients, and alemtuzumab-treated RRMS patients.

Sequence and reactivity tables for mature naïve B cells from HD18

|  | **HEAVY CHAIN** | | | | | | | **LIGHT CHAIN** | | | | | **REACTIVITY** | |
| --- | --- | --- | --- | --- | --- | --- | --- | --- | --- | --- | --- | --- | --- | --- |
| **mAb** | **VH** | **D** | **RF** | **JH** | **CDR3(aa)** | **Length** | **Mutations** | **Vk** | **Jk** | **CDR3(aa)** | **Length** | **Mutations** | **Poly** | **Hep-2** |
| **k-1** | 4-31 | 4-17 | 2 | 4 | CARADDYGEPFYW | 15 | 0 | 3-20 | 2 | CQQYGSSPMYTF | 12 | 0 | - | - |
| **k-5** | 4-34 | 4-11 | 2 | 4 | CARGRDIVETGPTELTNYDYYFDYW | 25 | 0 | 3-20 | 2 | CQQYGSSPPMYTF | 13 | 0 | - | + |
| **k-8** | 4-59 | 3-10 | 3 | 4 | CARGDTMVRGVPSSNDYW | 18 | 1 FR3 | 2-28 | 1 | CMQALQTPPTF | 11 | 0 | - | + |
| **k-11** | 1-18 | 3-16 | 1 | 4 | CARDNSGVVERFVVYYFDYW | 20 | 1 FR3 | 1-5 | 4 | CQQYNSYSSLTF | 12 | 1 FR3 | - | - |
| **k-14** | 3-23 | 3-22 | 2 | 4 | CANNYDSSGHIFGDYW | 16 | 1 FR1,1 FR3 | 1-6 | 4 | CLQDYNYPFTF | 11 | 0 | - | - |
| **k-18** | 3-48 | 3-10 | 1 | 5 | CARLPDRERFLAPW | 14 | 2 FR1 | 1-16 | 2 | CQHCNSYPYTF | 11 | 2 CDR3 | - | - |
| **k-20** | 3-9 | 3-22 | 2 | 4 | CAKDMSYYDSSGYYDYW | 17 | 0 | 1-39 | 2 | CQQSYSTLYTF | 11 | 1 CDR1 | - | - |
| **k-21** | 5-51 | 5-18 | 2 | 4 | CARHEGIQLSSRYFDYW | 17 | 0 | 3-20 | 1 | CQQYGSSPAWTF | 12 | 0 | - | - |
| **λ-2** | 4-34 | 5-24 | 3 | 4 | CARRRRDGYNLDYW | 14 | 0 | 1-51 | 2 | CGTWDSSLSATVF | 13 | 1 FR2 | - | - |
| **λ-6** | 4-34 | 6-13 | 3 | 2 | CASRAWQQLAKKNRKNIPKYYFDYW | 25 | 0 | 2-14 | 1 | CSSYTSSSTPYVF | 13 | 1 FR3 | + | + |
| **λ-7** | 3-23 | 3-16 | 2 | 3 | CAKGAQNYDYVWGSSFDIW | 19 | 1 FR1, 1 FR2 | 1-44 | 2 | CAAWDDSLNGPVF | 13 | 1 CDR1 | - | - |
| **λ-19** | 3-48 | 3-22 | 3 | 4 | CARAVVVVTSALYYFDYW | 18 | 0 | 3-21 | 2 | CQVWDSSSDHPLVVF | 15 | 0 | - | + |
| RF: reading frame; -: non-reactive; +: reactive; FR: framework; CDR: complementarity-determining region. | | | | | | | | | | | | | | |

Sequence and reactivity tables for mature naïve B cells from RRMS16

|  | **HEAVY CHAIN** | | | | | | | **LIGHT CHAIN** | | | | | **REACTIVITY** | |
| --- | --- | --- | --- | --- | --- | --- | --- | --- | --- | --- | --- | --- | --- | --- |
| **mAb** | **VH** | **D** | **RF** | **JH** | **CDR3(aa)** | **Length** | **Mutations** | **Vk** | **Jk** | **CDR3(aa)** | **Length** | **Mutations** | **Poly** | **Hep-2** |
| **k-A8B** | 4-4 | 3-10 | 3 | 2 | CASRPLGPYWYFDLW | 15 | 1 FR2 | 1-39 | 1 | CQQSYSTPWTF | 11 | 0 | - | + |
| **k-B10B** | 4-59 | 4-23 | 3 | 2 | CARDSGSTVVTPEYWHFDLW | 20 | 0 | 3-20 | 2 | CQQYGSSPMYTF | 12 | 0 | - | - |
| **k-C12A** | 3-30 | 3-9 | 2 | 5 | CARGLSSRPDLLTGYYYNWFDPW | 23 | 0 | 3-15 | 1 | CQQYNNWPPWTF | 12 | 0 | + | + |
| **k-D9B** | 3-48 | 1-26 | 1 | 4 | CARLVGATALW | 11 | 0 | 1-33 | 4 | CQQYDNLPPAF | 11 | 0 | - | + |
| **λ-C5A** | 3-43 | 3-3 | 2 | 5 | CAKDHSGADFWSGYSHRGFDPW | 22 | 0 | 1-51 | 3 | CGTWDSSLSAWVF | 13 | 1 FR1, 1 CDR2 | - | - |
| **λ-D1Γ** | 4-30 | 5-12 | 3 | 6 | CAREDVSGYGHYGMGVW | 17 | 1 CDR1 | 2-14 | 2 | CSSYTSSSPVVF | 12 | 0 | - | - |
| **λ-A5A** | 3-30 | 3-3 | 2 | 4 | CAKGDGAFWSGYSREDQNEDYW | 22 | 0 | 2-8 | 1 | CSSYAGSNRVF | 11 | 0 | - | - |
| **λ-A10A** | 1-69 | 3-10 | 1 | 4 | CARGSLGGFGEIDYW | 15 | 0 | 2-14 | 2 | CSSYTSSSPVVF | 12 | 0 | - | - |
| RF: reading frame; -: non-reactive; +: reactive; FR: framework; CDR: complementarity-determining region. | | | | | | | | | | | | | | |

Sequence and reactivity tables for mature naïve B cells from RRMS17

|  | **HEAVY CHAIN** | | | | | | | **LIGHT CHAIN** | | | | | **REACTIVITY** | |
| --- | --- | --- | --- | --- | --- | --- | --- | --- | --- | --- | --- | --- | --- | --- |
| **mAb** | **VH** | **D** | **RF** | **JH** | **CDR3(aa)** | **Length** | **Mutations** | **V** | **J** | **CDR3(aa)** | **Length** | **Mutations** | **Poly** | **Hep-2** |
| **k-Α9Γ** | 4-31 | 3-22 | 2 | 4 | CAISPTLYYYDGSGYFHW | 18 | 0 | 3-20 | 2 | CQQYGSSPMYTF | 12 | 1 FR1 | - | - |
| **k-Β11Α** | 3-30 | 3-22 | 2 | 4 | CARASYYYDSSGYYYWGYFDYW | 22 | 0 | 4-1 | 1 | CQQYYSTPTF | 10 | 1 FR3 | - | - |
| **k-Β12Α** | 3-30 | 5-12 | 3 | 4 | CAKPPWGSGYEYYFDYW | 17 | 0 | 1-16 | 4 | CQQYNSYPPTF | 11 | 0 | - | - |
| **λ-Α1Α** | 1-3 | 2-2 | 2 | 6 | CARDPVCSSTSCYGASNYYYYYGMDVW | 27 | 0 | 3-10 | 2 | CYSTDSSGKGVF | 12 | 0 | - | + |
| **λ-Α5Α** | 4-31 | 3-22 | 2 | 4 | CAISPTLYYYDSSGYFHW | 18 | 0 | 3-1 | 2 | CQAWDSSTAVVF | 13 | 1 FR3 | + | - |
| **λ-Α12Α** | 4-39 | 3-3 | 3 | 4 | CARLTIFGVVVXYW | 14 | 1 FR3 | 2-14 | 3 | CSSYTSSSPVVF | 12 | 1 FR3 | - | + |
| **λ-C9B** | 4-31 | 2-21 | 3 | 4 | CARGPPVVVTAMALDYW | 17 | 0 | 1-47 | 3 | CAAWDDSLSGPVF | 13 | 0 | - | + |
| **λ-D9A** | 1-46 | 2-2 | 3 | 4 | CARGPKPPEFDYW | 13 | 0 | 1-47 | 2 | CGTWDSSLSATVF | 13 | 1 FR2 | - | - |
| RF: reading frame; -: non-reactive; +: reactive; FR: framework; CDR: complementarity-determining region. | | | | | | | | | | | | | | |

Sequence and reactivity tables for mature naïve B cells from RRMS21

|  | **HEAVY CHAIN** | | | | | | | **LIGHT CHAIN** | | | | | **REACTIVITY** | |
| --- | --- | --- | --- | --- | --- | --- | --- | --- | --- | --- | --- | --- | --- | --- |
| **mAb** | **VH** | **D** | **RF** | **JH** | **CDR3(aa)** | **Length** | **Mutations** | **V** | **J** | **CDR3(aa)** | **Length** | **Mutations** | **Poly** | **Hep-2** |
| **k-B1Β** | 3-30 | 3-3 | 3 | 4 | CAKDPGIFGVVGDGYFDYW | 19 | 1 CDR2 | 4-1 | 3 | CATCACCTTC | 10 | 0 | - | - |
| **k-B3B** | 1-58 | 6-13 | 1 | 4 | CAAVYPLSSWYVLFDYW | 17 | 1 FR1 | 1-9 | 1 | CQQLNSYPPWTF | 12 | 0 | - | - |
| **k-D2Γ** | 1-24 | 6-13 | 2 | 5 | CATNRPRGIRVVRVFDPW | 18 | 0 | 4-1 | 1 | CQQYYSTPGTF | 11 | 0 | - | - |
| **k-A10A** | 4-59 | 6-6 | 1 | 5 | CARGYSSSSLGLEWVDPW | 18 | 0 | 1-33 | 4 | CQRYDNLPLTF | 11 | 1 CDR3 | - | - |
| **k-A12Γ** | 4-59 | 4-17 | 2 | 4 | CARGGVRDYGDYMPPFDYW | 19 | 1 CDR1 | 1-39 | 1 | CQQSYSTPWTF | 12 | 0 | - | - |
| **k-Β1Γ** | 3-21 | 2-21 | 2 | 4 | CARDRTDFYCGGDCGWFDPW | 20 | 0 | 3-20 | 1 | CQQYGSSPRAF | 12 | 0 | - | - |
| **k-Β5Γ** | 1-69 | 2-15 | 2 | 4 | CARDGPPNCSGGSCTFDYW | 19 | 0 | 3-11 | 3 | CQQRSNWPQFTF | 14 | 0 | - | - |
| **k-Β9Α** | 3-64 | 3-16 | 2 | 5 | CVKGRPHYDYIHNWFDPW | 18 | 0 | 1D-8 | 5 | CQQYYSFPITF | 11 | 0 | - | - |
| **k-ΗΒ12Α** | 3-23 | 5-12 | 1 | 4 | CAKALDQDIVAPGGW | 15 | 1 CDR1 | 3-11 | 4 | CQQRSNWPPALTF | 15 | 0 | + | + |
| **k-ΗC1Α** | 4-59 | 6-13 | 2 | 6 | CARAIAAAGYYYYYYMDVW | 19 | 0 | 1-39 | 2 | CQQSYSTPPYTF | 13 | 0 | - | - |
| **k-ΗC2Α** | 3-23 | 3-22 | 2 | 4 | CAKDYYYDSSGPFGYW | 16 | 0 | 1-8 | 3 | CQQYYSYPFTF | 12 | 0 | - | - |
| **k-ΗD7A** | 3-23 | 3-16 | 2 | 4 | CAKEGGSSDDYW | 12 | 0 | 1-NL1 | 2 | CQQYYSTPRTF | 12 | 0 | - | + |
| **λ-A8A** | 4-34 | 2-2 | 3 | 6 | CARGYIVVVPAAMPGGDYYMDVW | 24 | 0 | 2-8 | 1 | CSSYAGSNNLDF | 12 | 0 | - | + |
| RF: reading frame; -: non-reactive; +: reactive; FR: framework; CDR: complementarity-determining region. | | | | | | | | | | | | | | |

Sequence and reactivity tables for mature naïve B cells from ALE03

|  | **HEAVY CHAIN** | | | | | | | **LIGHT CHAIN** | | | | | **REACTIVITY** | |
| --- | --- | --- | --- | --- | --- | --- | --- | --- | --- | --- | --- | --- | --- | --- |
| **mAb** | **VH** | **D** | **RF** | **JH** | **CDR3(aa)** | **Length** | **Mutations** | **V** | **J** | **CDR3(aa)** | **Length** | **Mutations** | **Poly** | **Hep-2** |
| **k-A2B** | 3-11 | 5-18 | 1 | 4 | CAREEINHPTTNPNGDYW | 18 | 0 | 1-33 | 2 | CQQYDNLPRTF | 11 | 0 | - | - |
| **k-A8A** | 4-59 | 4-23 | 1 | 2 | CARGGTTVTSNWYFDLW | 14 | 0 | 1-33 | 4 | CQRYDNLPLTF | 11 | 1 FR2 | - | - |
| **k-Β11Α** | 4-59 | 3-9 | 1 | 3 | CARDFGLQGRYFDAEGAFDIW | 21 | 0 | 1D-39 | 1 | CQQSYSTRTF | 10 | 0 | - | + |
| **k-C5A** | 5-51 | 4-23 | 2 | 2 | CARRGDYGGTYWYFDLW | 17 | 0 | 3-15 | 1 | CQQYNNRPSWTF | 12 | 0 | - | - |
| **k-C8A** | 4-30 | 3-10 | 1 | 6 | CARDVLWFGELSWYAWPDVW | 20 | 0 | 1-9 | 1 | CQQLNSYPPWTF | 11 | 0 | - | - |
| **k-Α7Γ** | 3-30-3 | 3-10 | 1 | 4 | CARDFFPPWFGSNYRFDYW | 19 | 1 CDR2 | 2-30 | 1 | CMQGTHWTWTF | 11 | 0 | + | + |
| **k-C7B** | 3-21 | 2-15 | 2 | 5 | CARYCSGGSCLGGFDPW | 17 | 0 | 3-15 | 2 | CQQYNNWPPYTF | 11 | 0 | - | + |
| **k-C2A** | 3-11 | 6-19 | 2 | 4 | CARGRLGLIAVAGPFEDYW | 19 | 0 | 1-8 | 1 | CQQYYSYPQTF | 11 | 0 | - | - |
| RF: reading frame; -: non-reactive; +: reactive; FR: framework; CDR: complementarity-determining region. | | | | | | | | | | | | | | |

Sequence and reactivity tables for mature naïve B cells from ALE25

|  | **HEAVY CHAIN** | | | | | | | **LIGHT CHAIN** | | | | | **REACTIVITY** | |
| --- | --- | --- | --- | --- | --- | --- | --- | --- | --- | --- | --- | --- | --- | --- |
| **mAb** | **VH** | **D** | **RF** | **JH** | **CDR3(aa)** | **Length** | **Mutations** | **V** | **J** | **CDR3(aa)** | **Length** | **Mutations** | **Poly** | **Hep-2** |
| **k-A4B** | 3-9 | 3-10 | 3 | 5 | CAKDGAVRGVYYGSGWFDPW | 20 | 0 | 1-5 | 3 | CQQYNSYS#FTF | 12 | 1 CDR3 | - | - |
| **k-A6B** | 4-34 | 6-6 | 3 | 4 | CARGRWSSSSLCDYW | 15 | 1 FR3 | 3-15 | 2 | CQQYNNWPPYTF | 13 | 0 | - | - |
| **k-Α8Α** | 1-69 | 6-13 | 1 | 6 | CAREDSSSLRRGYYYGMDVW | 20 | 0 | 4-1 | 4 | CQQYYSTPLTF | 11 | 0 | - | - |
| **k-B1A** | 4-59 | 6-6 | 2 | 4 | CARDSSIAARPGAGFDYW | 18 | 0 | 1-33 | 4 | CQQYDNLPLTF | 11 | 0 | - | - |
| **k-B4Γ** | 4-30-4 | 2-2 | 1 | 3 | CARVHQLLRSNYYDPTPTDAFDIW | 24 | 1 CDR2, 1 FR3 | 3-20 | 2 | CQQYGSSPYTF | 11 | 0 | - | - |
| **k-C2A** | 4-34 | 2-2 | 1 | 6 | CARGASPRPYYYYGMDVW | 18 | 0 | 1-39 | 4 | CQQSYSTPLTF | 11 | 0 | - | - |
| **k-C4A** | 4-4 | 6-13 | 2 | 1 | CARDSQTIAAAGAFRDW | 17 | 1 FR2, 1 FR3 | 3-20 | 4 | CQQYGSSPLTF | 11 | 1 FR2 | - | - |
| **k-D7A** | 3-48 | 2-21 | 2 | 5 | CAREPSPYCGGDCYALSNWGFDPW | 24 | 1 CDR1 | 3-20 | 2 | CQQYGSSPTYTF | 12 | 0 | - | - |
| **k-Α12Β** | 1-2 | 2-8 | 2 | 4 | CACTVGVGYW | 10 | 1 CDR2 | 1D-8 | 4 | CQQYYSFPLTF | 11 | 0 | + | + |
| **k-C7Γ** | 3-23 | 6-13 | 1 | 1 | CAKENGYSSSWSVEAYFQHR | 20 | 0 | 4-1 | 1 | CQQYYSTPRTF | 11 | 0 | - | - |
| **k-C9A** | 3-11 | 2-8 | 1 | 5 | CASHRTYYYGSALVW | 15 | 0 | 1-39 | 1 | CQQSYSTPWTF | 11 | 0 | - | - |
| **k-C11Δ** | 1-69 | 6-6 | 1 | 5 | CARGEDSSSSGWFDPW | 16 | 0 | 3-20 | 1 | CQQYGSSPRTF | 11 | 0 | - | - |
| RF: reading frame; -: non-reactive; +: reactive; FR: framework; CDR: complementarity-determining region. | | | | | | | | | | | | | | |

Sequence and reactivity tables for mature naïve B cells from ALE27

|  | **HEAVY CHAIN** | | | | | | | **LIGHT CHAIN** | | | | | **REACTIVITY** | |
| --- | --- | --- | --- | --- | --- | --- | --- | --- | --- | --- | --- | --- | --- | --- |
| **mAb** | **VH** | **D** | **RF** | **JH** | **CDR3(aa)** | **Length** | **Mutations** | **V** | **J** | **CDR3(aa)** | **Length** | **Mutations** | **Poly** | **Hep-2** |
| **k-A6A** | 3-23 | 4-17 | 2 | 4 | CAKGAHDYGDQGLVLFDYW | 20 | 0 | 3-15 | 1 | CQQYNNWPPTF | 11 | 0 | + | + |
| **k-A8Α** | 3-30 | 4-17 | 2 | 2 | CAKDLGDYVEVFDLW | 15 | 0 | 3-15 | 2 | CQQYNNXPYTF | 11 | 1 CDR3 | - | - |
| **k-Α11Γ** | 5-51 | 2-15 | 3 | 2 | CARPEGVVAATPWYFDLW | 18 | 0 | 2-28 | 4 | CMQALQTRTF | 10 | 0 | - | - |
| **k-B10Δ** | 1-2 | 1-14 | 1 | 4 | CARDLNGWQPDYW | 13 | 1 FR2 | 3-15 | 2 | CQQYNNWPPYTF | 12 | 1 FR2 | - | - |
| **k-D3B** | 3-33 | 3-33 | 1 | 4 | CASDGAAGWIVGATDIDYW | 19 | 1 CDR3 | 1-39 | 2 | CQQSYSTLYTF | 11 | 0 | - | - |
| **k-D10Δ** | 3-30 | 2-21 | 2 | 6 | CAKDRGGDYGSYYYGMDVW | 19 | 1 FR3 | 3-20 | 2 | CQQYGSPPMYTF | 12 | 1 FR3 | - | - |
| **λ-Α10Α** | 3-9 | 3-22 | 2 | 4 | CAKDIQETPTYYYDSSGYFDYW | 22 | 0 | 1-40 | 2 | CQSYDSSLSGFVVF | 14 | 0 | - | - |
| RF: reading frame; -: non-reactive; +: reactive; FR: framework; CDR: complementarity-determining region. | | | | | | | | | | | | | | |

Sequence and reactivity tables for mature naïve B cells from ALE40

|  | **HEAVY CHAIN** | | | | | | | **LIGHT CHAIN** | | | | | **REACTIVITY** | |
| --- | --- | --- | --- | --- | --- | --- | --- | --- | --- | --- | --- | --- | --- | --- |
| **mAb** | **VH** | **D** | **RF** | **JH** | **CDR3(aa)** | **Length** | **Mutations** | **V** | **J** | **CDR3(aa)** | **Length** | **Mutations** | **Poly** | **Hep-2** |
| **k-A1A** | 3-21 | 1-1 | 1 | 1 | CASGTGTTGYW | 11 | 0 | 3-11 | 1 | CQQRSNWPPTF | 11 | 0 | - | + |
| **k-Β8Α** | 1-46 | 1-26 | 1 | 4 | CARDGIVGATDAFDIW | 16 | 1 FR2, 1 FR3 | 1-39 | 4 | CQQSYSTPLTF | 11 | 0 | - | - |
| **k-C7B** | 4-39 | 1-26 | 3 | 4 | CARLRYSGSYMDYW | 14 | 0 | 4-1 | 1 | CQQYYSTPGTF | 11 | 1 FR2 | + | + |
| **k-C10A** | 1-3 | 3-3 | 2 | 6 | CASLYYDFWSGYYHDRAGDYYGMDVW | 26 | 0 | 3-15 | 2 | CQQYNNWPPYTF | 12 | 0 | - | - |
| **k-D5A** | 3-33 | 3-9 | 2 | 4 | CARGADILTGYYGPDYW | 17 | 1 FR2 | 1-6 | 1 | CLQDYNYPRTF | 11 | 0 | - | - |
| **k-D9B** | 5-10-1 | 2-15 | 2 | 3 | CARLEVCDGSCYIAAFDIW | 19 | 1 FR3 | 3-20 | 2 | CQQYGSSPLYTF | 12 | 0 | - | - |
| **k-B3** | 1-3 | 6-13 | 2 | 3 | CARTNIAAAGNVW | 13 | 0 | 3-11 | 1 | CQQRSNWPPTF | 11 | 0 | - | + |
| **k-B7** | 1-69 | 3-10 | 2 | 5 | CASESGRGWFDPW | 13 | 1 CDR3 | 3-20 | 2 | CQQYGSSLYTF | 11 | 0 | - | - |
| **k-B5 _(2nd cDNAplate)_** | 1-69 | 2-2 | 2 | 4 | CARRYCSSTSCYLNYW | 16 | 1 FR2 | 1-5 | 2 | CQQYNSYSETF | 11 | 0 | - | + |
| **λ-B5A** | 5-51 | 1-14 | 3 | 2 | CARTRHYYYYYGMDVW | 16 | 0 | 3-21 | 1 | CQVWDSSSDHPGVF | 14 | 0 | - | - |
| **λ-C7** | 3-21 | 5-12 | 1 | 6 | CARADGGAIDYYYYGMDVW | 19 | 0 | 3-21 | 3 | CQVWDSSSDHQGVF | 14 | 0 | - | - |
| RF: reading frame; -: non-reactive; +: reactive; FR: framework; CDR: complementarity-determining region. | | | | | | | | | | | | | | |

Sequence and reactivity tables for mature naïve B cells from ALE 33

|  | **HEAVY CHAIN** | | | | | | | **LIGHT CHAIN** | | | | | **REACTIVITY** | |
| --- | --- | --- | --- | --- | --- | --- | --- | --- | --- | --- | --- | --- | --- | --- |
| **mAb** | **VH** | **D** | **RF** | **JH** | **CDR3(aa)** | **Length** | **Mutations** | **V** | **J** | **CDR3(aa)** | **Length** | **Mutations** | **Poly** | **Hep-2** |
| **k-A3A** | 3-43 | 3-22 | 2 | 4 | CAKDKAYDSSGYYFDYW | 17 | 0 | 1-12 | 4 | CQQANSXPLLTF | 12 | 1 CDR3 | - | - |
| **k-A6A** | 3-23 | 2-15 | 2 | 4 | CAKDQGYCSGGSCSFFDYW | 19 | 0 | 1-12 | 3 | CQQANXFPFTF | 11 | 0 | - | + |
| **k-B10A** | 4-39 | 6-19 | 1 | 2 | CARRVDSAGWGWYFDLW | 17 | 0 | 1-39 | 2 | CQQSYSTPYTF | 11 | 0 | + | + |
| **k-C3A** | 1-2 | 6-6 | 2 | 5 | CATSNAAEENSNWFDPW | 17 | 0 | 3-20 | 2 | CQQYGSSPYTF | 11 | 1 FR1 | - | - |
| **k-D6A** | 3-48 | 1-26 | 1 | 4 | CARYLLVGATAGIDYW | 16 | 0 | 1-12 | 4 | CQQANSFPITF | 11 | 0 | - | - |
| **k-A4** | 4-61 | 3-10 | 2 | 6 | CARGGSYYYYGMDVW | 15 | 1 CDR1 | 1-12 | 1 | CQQANSFPRTF | 11 | 0 | + | + |
| **λ-A4A** | 5-51 | 3-10 | 2 | 4 | CARHGYYGSGSYYNFDYW | 18 | 0 | 1-40 | 3 | CQSYDSSLSGWVF | 13 | 0 | - | - |
| **λ-A5A** | 4-39 | 3-22 | 1 | 4 | CARQGKYYFDYW | 12 | 0 | 3-1 | 1 | CQAWDSSTGVF | 11 | 0 | - | - |
| **λ-D12A** | 4-34 | 2-8 | 2 | 4 | CARENCTNGVCYVGQEGIDYW | 21 | 0 | 3-25 | 3 | CQSADSSGTYRVF | 13 | 0 | - | + |
| **λ-Α9** | 3-23 | 6-19 | 1 | 4 | CAKDYYSSGWFAPGFDYW | 18 | 0 | 2-23 | 2 | CCSYAGSSTFVVF | 13 | 0 | + | + |
| **λ-B7** | 4-39 | 1-26 | 1 | 4 | CACVPLSYASIVGATPFDYW | 20 | 0 | 1-40 | 2 | CQSYDSSLSAVVX | 13 | 0 | - | + |
| **Λ-Β8** | 4-34 | 3 | 2 | 4 | CARDTVTYVWGSYRPKWTFDYW | 22 | 0 | 2-23 | 2 | CCSYAGSVVF | 10 | 1 FR3 | + | + |
| RF: reading frame; -: non-reactive; +: reactive; FR: framework; CDR: complementarity-determining region. | | | | | | | | | | | | | | |

Sequence and reactivity tables for mature naïve B cells from ALE 46

|  | **HEAVY CHAIN** | | | | | | | **LIGHT CHAIN** | | | | | **REACTIVITY** | |
| --- | --- | --- | --- | --- | --- | --- | --- | --- | --- | --- | --- | --- | --- | --- |
| **mAb** | **VH** | **D** | **RF** | **JH** | **CDR3(aa)** | **Length** | **Mutations** | **V** | **J** | **CDR3(aa)** | **Length** | **Mutations** | **Poly** | **Hep-2** |
| **k-B2B** | 3-9 | 2-21 | 3 | 4 | CAKDVHREVVGTLDYW | 16 | 0 | 1-27 | 1 | CQKYNXAPTWTF | 12 | 1 CDR3 | - | - |
| **k-B6A** | 5-51 | 2-2 | 3 | 4 | CARQSPHAFDYW | 12 | 0 | 1-27 | 4 | CQKYNSAPLTF | 11 | 0 | - | - |
| **k-B8B** | 3-33 | 2-15 | 2 | 4 | CARDTGYCSGGSCPILDYW | 19 | 0 | 1-33 | 3 | CQQYDNLLRGVTF | 13 | 1 CDR1 | - | - |
| **k-Β10Δ** | 3-43 | 3-22 | 2 | 4 | CAKDYYPGRDSSGYYNW | 17 | 0 | 1-39 | 4 | CQQSYSTP#LTF | 12 | 0 | - | - |
| **k-C4A** | 1-2 | 1-26 | 1 | 4 | CASLGTEDFDYW | 12 | 0 | 1-5 | 2 | CQQYNSNSRXT | 11 | 0 | - | - |
| **k-B6** | 3-9 | 4-17 | 2 | 4 | CAKDYGDW | 8 | 1 FR1 | 3-11 | 4 | CQQRSNWPPGLTF | 13 | 1 CDR2 | - | - |
| **k-B7** | 3-30 | 1-26 | 2 | 4 | CAREWWELHQFDYW | 14 | 0 | 1-17 | 1 | CLQHNSYPWTF | 11 | 1 FR2 | + | + |
| **k-B12** | 1-46 | 6-6 | 2 | 4 | CARDQIAARPSNFDYW | 16 | 0 | 1-39 | 2 | CQQSYXTPPCTF | 12 | 0 | - | + |
| **λ-A4** | 4-4 | 3-3 | 2 | 6 | CARDRGYDFRGMDVW | 15 | 1 CDR1 | 3-25 | 3 | CQSADSSGTYWVF | 13 | 0 | + | - |
| **λ-A5** | 1-24 | 3-3 | 2 | 4 | CAITHRAYYDFWSGYFRFDYW | 21 | 1 CDR1 | 3-25 | 3 | CQSADSSGTYWVF | 13 | 0 | - | + |
| **λ-B3** | 1-69 | 3-22 | 2 | 5 | CARVHGDYYDSSGENSPFDPW | 21 | 1 FR2 | 1-47 | 3 | CAAWDDSLSA#WVF | 14 | 0 | - | - |
| **λ-A5** | 5-51 | 5-18 | 1 | 5 | CARLGSTPNMNWFDPW | 16 | 0 | 1-51 | 5 | CGTWDSSLSAVVF | 13 | 0 | - | + |
| **λ-A8** | 3-21 | 4-23 | 2 | 1 | CAREGNLSSW | 10 | 0 | 1-51 | 2 | CGTWDSSLSAVVX | 13 | 0 | - | - |
| **λ-A12** | 3-23 | 3-22 | 2 | 6 | CAKESSSSGWYEEGGYWYFDLW | 22 | 0 | 1-40 | 2 | CCSYAGSSTFVVF | 13 | 0 | - | - |
| RF: reading frame; -: non-reactive; +: reactive; FR: framework; CDR: complementarity-determining region. | | | | | | | | | | | | | | |

**Supplementary Table 4.** TSHR cell-based assay (CBA) results for HD, RRMS, alemtuzumab-treated RRMS patients

| **Samples** | **rIgG** | **ΔMFI** |
| --- | --- | --- |
| **Positive control** | M22 | 13.043 |
| **Negative control** | PBS | >0.0 |
| **HD18** | H1 | >0.0 |
|  | H2 | >0.0 |
|  | H5 | >0.0 |
|  | H6 | >0.0 |
|  | H7 | >0.0 |
|  | H8 | >0.0 |
|  | H14 | >0.0 |
|  | H21 | >0.0 |
|  | H18 | >0.0 |
|  | H19 | >0.0 |
|  | H20 | >0.0 |
|  | H10 | >0.0 |
| **RRMS17** | HA9 | >0.0 |
|  | HA5 | >0.0 |
|  | HA12 | >0.0 |
|  | HB12 | >0.0 |
|  | HC9 | >0.0 |
|  | HA1 | >0.0 |
|  | HB11 | >0.0 |
|  | HD9 | >0.0 |
| **RRMS16** | ΗΒ10 | >0.0 |
|  | ΗC5 | >0.0 |
|  | HC12 | >0.0 |
|  | HD1 | >0.0 |
|  | HD9 | >0.0 |
|  | HA5 | >0.0 |
|  | HA10 | >0.0 |
|  | HA8 | >0.0 |
| **RRMS21** | HB1 | >0.0 |
|  | HB3 | >0.0 |
|  | HD2 | >0.0 |
|  | HA10 | >0.0 |
|  | HA12 | >0.0 |
|  | HB1 | >0.0 |
|  | HB5 | >0.0 |
|  | HB9 | >0.0 |
|  | HB12 | >0.0 |
|  | HC1 | >0.0 |
|  | HC2 | >0.0 |
|  | HA8 | >0.0 |
|  | HD7 | >0.0 |
| **ALE03** | HA2 | >0.0 |
|  | HA8 | >0.0 |
|  | HB11 | >0.0 |
|  | HC5 | >0.0 |
|  | HC8 | >0.0 |
|  | HA7 | >0.0 |
|  | HC7 | >0.0 |
|  | HC2 | >0.0 |
| **ALE40** | A1 | >0.0 |
|  | B8 | >0.0 |
|  | C7 | >0.0 |
|  | C10 | >0.0 |
|  | D5 | >0.0 |
|  | D9 | >0.0 |
|  | B3 | >0.0 |
|  | B7 | >0.0 |
|  | B5 | >0.0 |
|  | B5A | >0.0 |
|  | C7 | >0.0 |
| **ALE 33** | A3 | >0.0 |
|  | B10 | >0.0 |
|  | C3 | >0.0 |
|  | C5 | >0.0 |
|  | C11 | >0.0 |
|  | D6 | >0.0 |
|  | A4 | >0.0 |
|  | A5 | >0.0 |
|  | D12 | >0.0 |
|  | A6 | >0.0 |
| **ALE 46** | B2 | >0.0 |
|  | B6 | >0.0 |
|  | B8 | >0.0 |
|  | B10 | >0.0 |
|  | C4 | >0.0 |
|  | A4 | >0.0 |
|  | A5 | >0.0 |
|  | B3 | >0.0 |
| **ALE 25** | A6 | >0.0 |
|  | B1 | >0.0 |
|  | A8 | >0.0 |
|  | B4 | >0.0 |
|  | C2 | >0.0 |
|  | C4 | >0.0 |
|  | C7 | >0.0 |
|  | D7 | >0.0 |
|  | A12 | >0.0 |
|  | C9 | >0.0 |
|  | C11 | >0.0 |
|  | A4 | >0.0 |
| **ALE 27** | A6 | >0.0 |
|  | A8 | >0.0 |
|  | A11 | >0.0 |
|  | B1 | >0.0 |
|  | B1O | >0.0 |
|  | D3 | >0.0 |
|  | D10 | >0.0 |
|  | A10 | >0.0 |
|  | C6 | >0.0 |
| ALE: Alemtuzumab-treated RRMS patients; HD: Healthy donor; RRMS: Relapsing-remitting multiple sclerosis | | |

**Supplementary Table 5.** BCR sorting into naive and memory sequences based on mutation counts for samples

| **Samples** | **Total productive sequences** | **Naïve B cells sequences** | **% Naïve B cells sequences** | **Memory B cells sequences** | **% Memory B cells sequences** |
| --- | --- | --- | --- | --- | --- |
| HD 13 | 58403 | 13699 | 23.4 | 44704 | 76.6 |
| HD18 | 27228 | 6423 | 23.6 | 20805 | 76.4 |
| HD19 | 7002 | 2608 | 37.2 | 4394 | 62.8 |
| RRMS 09 | 2578 | 650 | 25.2 | 1928 | 74.8 |
| RRMS 16 | 22854 | 7909 | 34.6 | 14925 | 65.4 |
| RRMS 17 | 12484 | 4008 | 32.1 | 8476 | 67.9 |
| RRMS 21 | 4981 | 1014 | 20.4 | 3967 | 79.6 |
| ALE 03 | 19306 | 11307 | 58.6 | 7999 | 41.4 |
| ALE 24 | 3740 | 1559 | 41.7 | 2191 | 58.3 |
| ALE 25 | 5720 | 3023 | 52.8 | 2697 | 47.2 |
| ALE26 | 19504 | 11189 | 57.4 | 8315 | 42.6 |
| ALE 27 | 36292 | 8055 | 22.2 | 28237 | 77.8 |
| ALE 29 | 3249 | 1614 | 49.7 | 1635 | 50.3 |
| ALE 40 | 3610 | 1665 | 46.1 | 1945 | 53.9 |
| ALE: Alemtuzumab-treated RRMS patients; HD: Healthy donor; RRMS: Relapsing-remitting multiple sclerosis | | | | | |

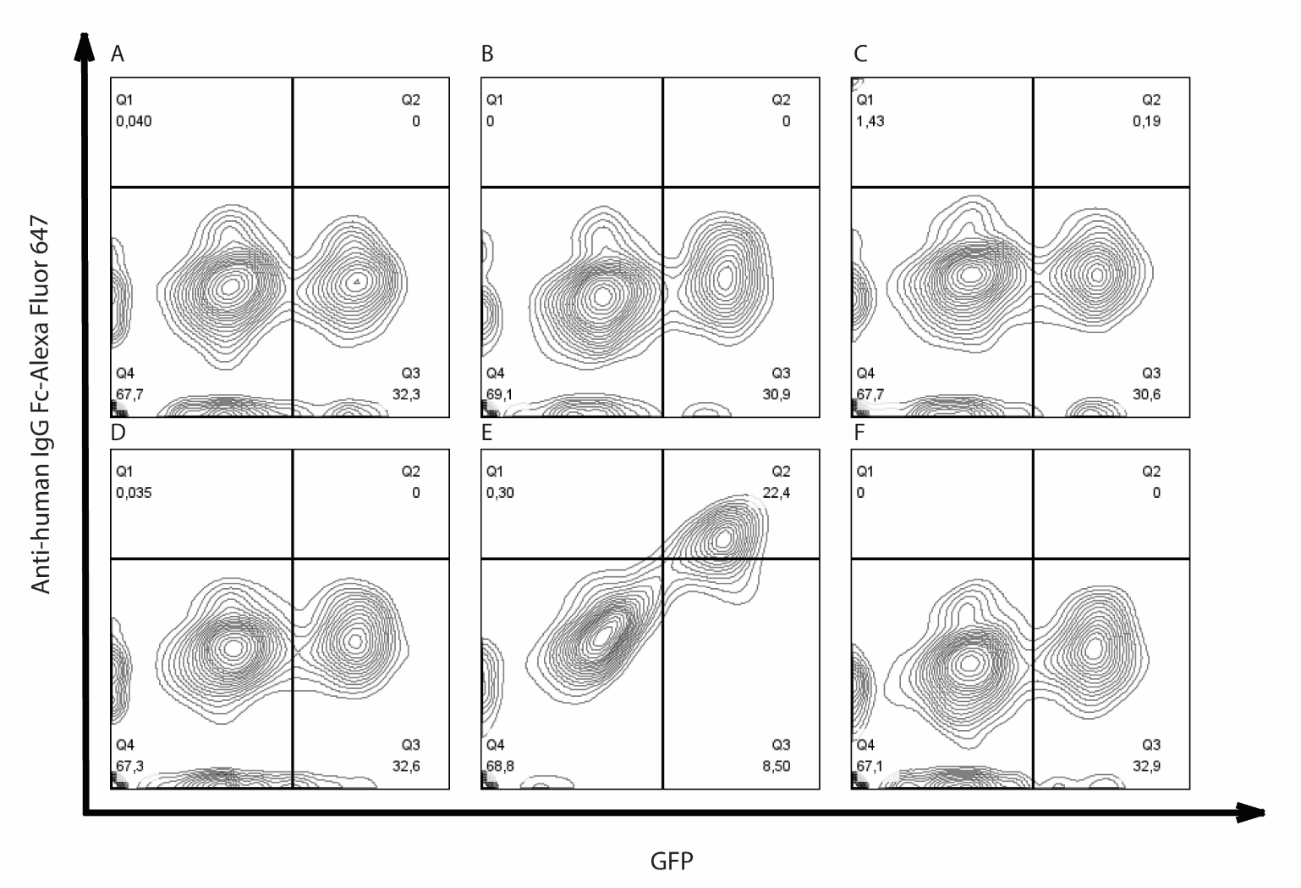

**Supplementary Figure 1.** Representative TSHR cell-based assay (CBA) flow cytometry plots. Purified mature naïve B cell-derived recombinant antibodies from HDs, RRMS, and alemtuzumab-treated specimens were tested for surface binding to TSHR on TSHR-GFP-transfected HEK cells, M22 a TSHR autoantibody was used as a positive control; PBS was used as a negative control. All antibodies were tested in duplicate at a concentration of 2μg/mL in 1% BSA in PBS. The x-axis represents GFP fluorescence intensity; the y-axis represents Alexa Fluor 647 fluorescence intensity. Cells in the upper right quadrant are transfected with TSHR-GFP and bind TSHR antibodies. **(A)** mAb HA12 from ALE25, positive for both polyreactivity and autoreactivity; **(B)** mAb A4B from ALE25, negative for both polyreactivity and autoreactivity; **(C)** mAb HB10 from ALE46, positive for polyreactivity, negative for autoreactivity; **(D)** mAb HA4 from ALE46, negative for polyreactivity, positive for autoreactivity; **(E)** M22, positive control^1^; **(F)** PBS, negative control. Ale: Alemtuzumab-treated RRMS patients; BSA: Bovine Serum Albumin; GFP: Green Fluorescent Protein; HD: Healthy donor; HEK: Human Embryonic Kidney; LPS: Lipopolysaccharide; mAb: Monoclonal antibody; PBS: Phosphate-Buffered Saline; RRMS: Relapsing-remitting multiple sclerosis; TSHR: Thyroid-stimulating hormone receptor.

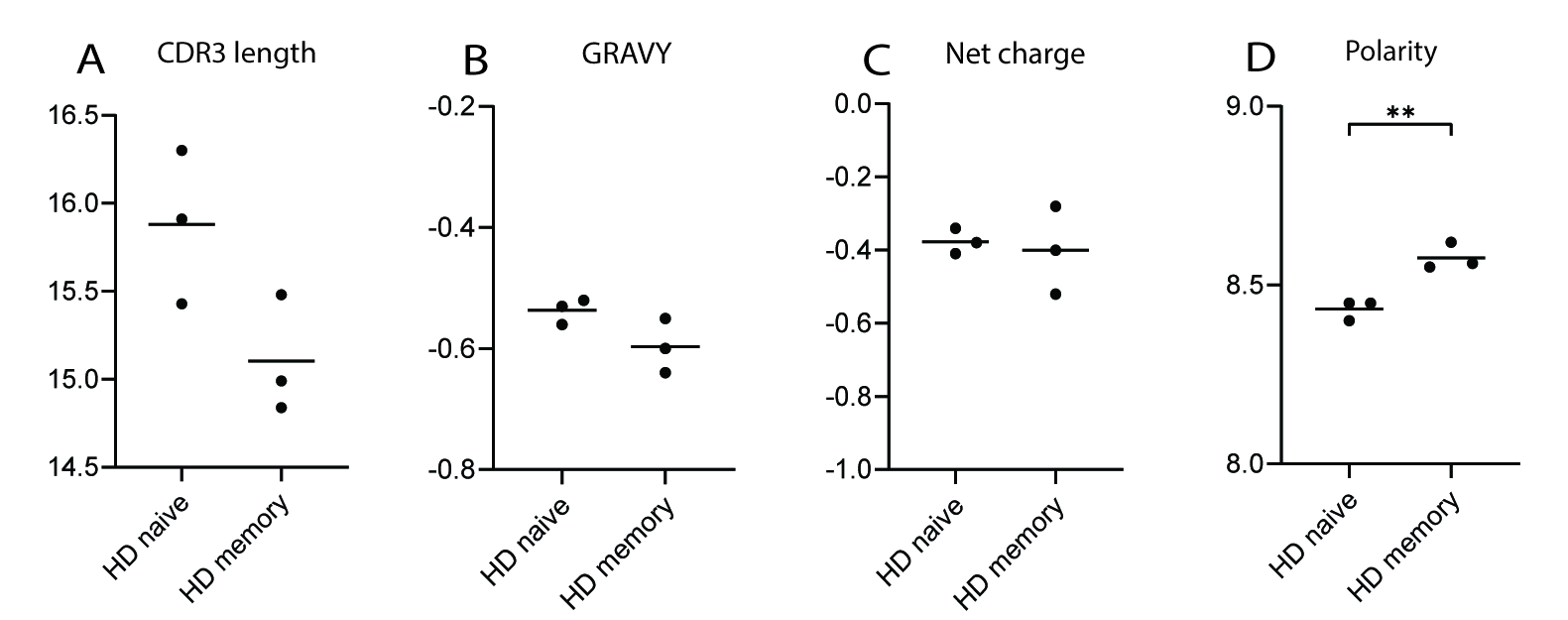

**Supplementary Figure 2.** IgH CDR3 property comparisons between naïve and memory B cells of HDs. (A) CDR3 length, (B) grand average hydropathy index (GRAVY), (C) net charge, (D) polarity (P = 0.007). Mann-Whitney testing was used for the assessment of differences between groups, and p values are indicated if significant (**P ≤ 0.01; *P ≤ 0.05). CDR3: complementarity-determining region 3; HD: Healthy donor; IgH: Immunoglobulin heavy chain.

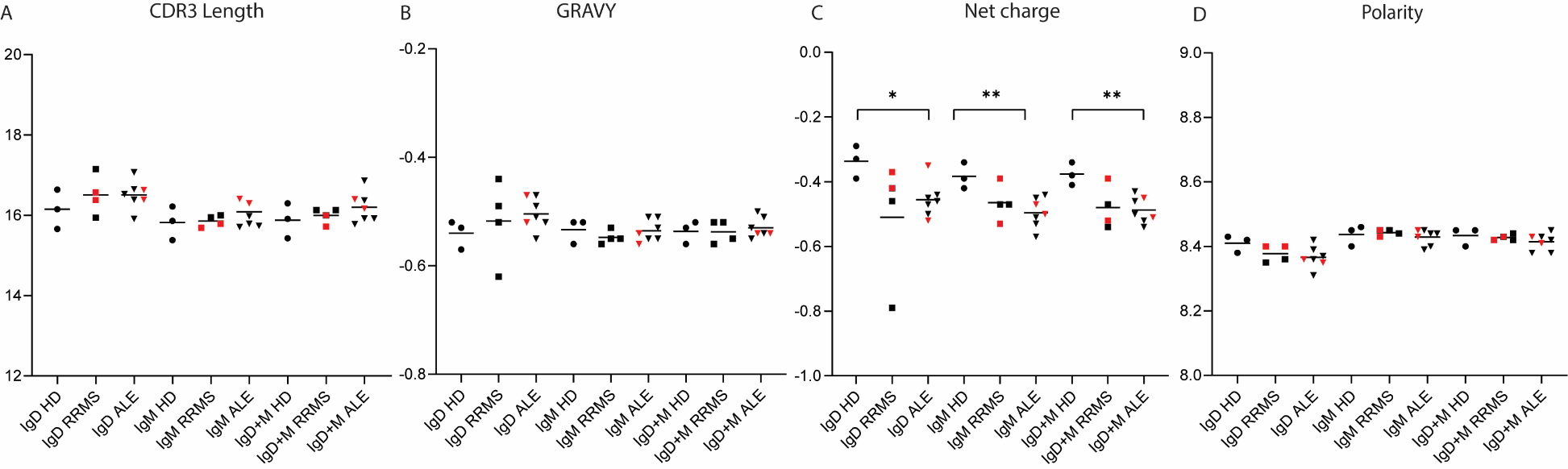

**Supplementary Figure 3.** Analysis of naïve IgH CDR3 properties highlighting patients with a high fraction of autoreactive naïve B cells. The following properties were calculated per Ig class: **(A)** CDR3 length, **(B)** the grand average hydropathy index (GRAVY), **(C)** net charge, and **(D)** polarity. Black circles represent individual HD values, black squares those of immunotherapy-naïve RRMS patients and black triangles those of alemtuzumab-treated RRMS patients. Among the two latter, red symbols represent patients with a compromised peripheral tolerance checkpoint as determined by mAb autoreactivity ELISA. Differences between groups (HD, immunotherapy-naïve RRMS, and alemtuzumab-treated RRMS) were tested with the Mann-Whitney test, and p values are indicated if significant (**P ≤ 0.01; *P ≤ 0.05). ALE: Alemtuzumab-treated RRMS patients; CDR: Complementarity-Determining Region; HD: Healthy donor; Ig: immunoglobulin; IgH: heavy chain; NGS: Next-generation sequencing; RRMS: Relapsing-remitting multiple sclerosis.

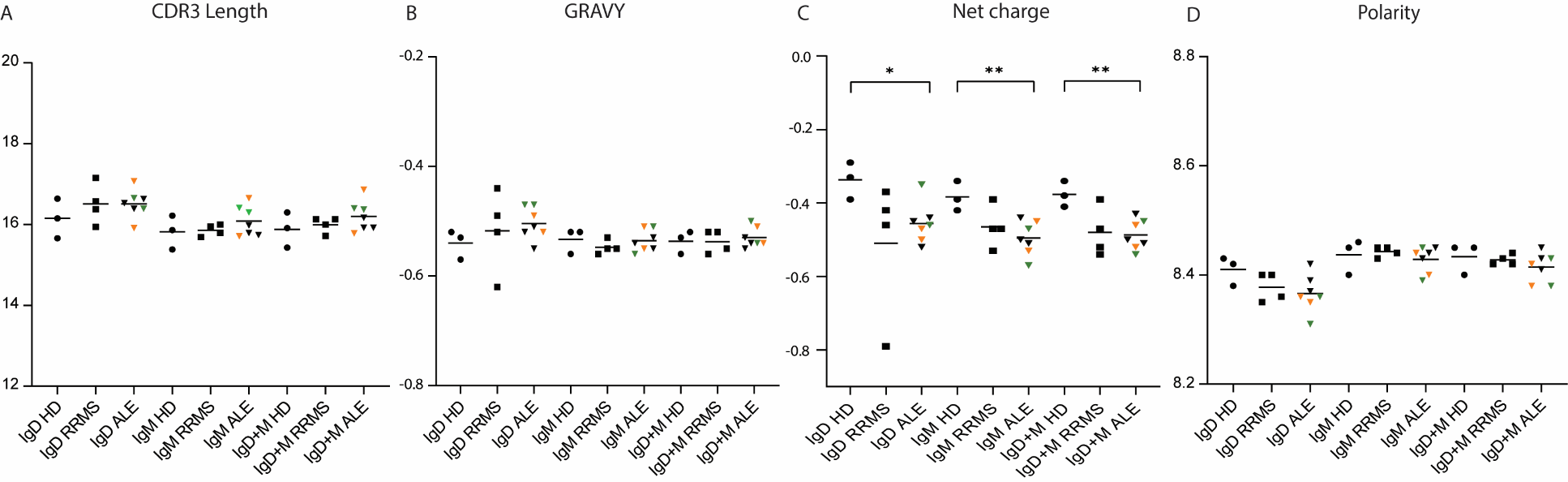

**Supplementary Figure 4.** Analysis of naïve IgH CDR3 properties highlighting MS activity/secondary autoimmunity up to the timepoint of sampling. The following properties were calculated per Ig class: (A) CDR3 length, (B) the grand average hydropathy index (GRAVY), (C) net charge, and (D) polarity. Black circles represent individual HD values, black squares those of immunotherapy-naïve RRMS patients and triangles those of alemtuzumab-treated RRMS patients. Among the latter, green triangles represent alemtuzumab-treated patients without signs of MS activity and without secondary autoimmunity, orange triangles alemtuzumab-treated patients with both signs of MS activity and of secondary autoimmunity, and black triangles alemtuzumab-treated patients without MS activity but with secondary autoimmunity. Differences between groups (HD, immunotherapy-naïve RRMS, and alemtuzumab treated RRMS) were tested with the Mann-Whitney test, and p values are indicated when significant (**P ≤ 0.01; *P ≤ 0.05). ALE: Alemtuzumab-treated RRMS patients; CDR: Complementarity-Determining Region; HD: Healthy donor; Ig: immunoglobulin; IgH: heavy chain; NGS: Next-generation sequencing; RRMS: Relapsing-remitting multiple sclerosis.
